# The Complexity of Tobacco Smoke-Induced Mutagenesis in Head and Neck Cancer

**DOI:** 10.1101/2024.04.15.24305006

**Authors:** Laura Torrens, Sarah Moody, Ana Carolina de Carvalho, Mariya Kazachkova, Behnoush Abedi-Ardekani, Saamin Cheema, Sergey Senkin, Thomas Cattiaux, Ricardo Cortez Cardoso Penha, Joshua R. Atkins, Valérie Gaborieau, Priscilia Chopard, Christine Carreira, Ammal Abbasi, Erik N. Bergstrom, Raviteja Vangara, Jingwei Wang, Stephen Fitzgerald, Calli Latimer, Marcos Diaz-Gay, David Jones, Jon Teague, Felipe Ribeiro Pinto, Luiz Paulo Kowalski, Jerry Polesel, Fabiola Giudici, José Carlos de Oliveira, Pagona Lagiou, Areti Lagiou, Marta Vilensky, Dana Mates, Ioan N. Mates, Lidia M. Arantes, Rui Reis, Jose Roberto V. Podesta, Sandra V. von Zeidler, Ivana Holcatova, Maria Paula Curado, Cristina Canova, Elenora Fabianova, Paula A. Rodríguez-Urrego, Laura Humphreys, Ludmil B. Alexandrov, Paul Brennan, Michael R. Stratton, Sandra Perdomo

## Abstract

Tobacco smoke, alone or combined with alcohol, is the predominant cause of head and neck cancer (HNC). Here, we further explore how tobacco exposure contributes to cancer development by mutational signature analysis of 265 whole-genome sequenced HNC from eight countries. Six tobacco-associated mutational signatures were detected, including some not previously reported. Differences in HNC incidence between countries corresponded with differences in mutation burdens of tobacco-associated signatures, consistent with the dominant role of tobacco in HNC causation. Differences were found in the burden of tobacco-associated signatures between anatomical subsites, suggesting that tissue-specific factors modulate mutagenesis. We identified an association between tobacco smoking and three additional alcohol-related signatures indicating synergism between the two exposures. Tobacco smoking was associated with differences in the mutational spectra and repertoire of driver mutations in cancer genes, and in patterns of copy number change. Together, the results demonstrate the multiple pathways by which tobacco smoke can influence the evolution of cancer cell clones.

## INTRODUCTION

Head and neck cancer (HNC), including malignancies affecting the mouth, pharynx, and larynx, represents ∼4% of the global cancer burden, with an annual incidence of about 750,000 new cases^1^. The incidence rate of HNC varies between different countries, largely reflecting the distribution of its main risk factors including tobacco smoking, alcohol consumption^1,2^, and infection with high-risk strains of human papillomavirus (HPV) for oropharynx cancer^3–5^. Other proposed risk factors include consumption of hot beverages, obesity, and poor oral health, although evidence for their role in HNC is limited^6–8^. In addition, a substantial proportion of head and neck cancers (about 42% for women and 26% for men) cannot be attributed to known lifestyle habits or exposures^9^.

Epidemiological studies in Europe and America suggest that seven out of 10 HNC cancers are caused by preventable behavioral risk factors, with tobacco use, either alone or in combination with alcohol, accounting for most cases^9^. Conversely, alcohol use on its own is responsible for only ∼4% of the disease burden, suggesting a limited impact on HNC burden. This raises the question of whether alcohol acts as an independent carcinogen or simply enhances the known carcinogenic effect of tobacco. Furthermore, the susceptibility to these exposures varies depending on the anatomical region, with smoking posing a higher risk for developing larynx cancer and the risk associated with alcohol being greater for other subsites^10^.

Considering the dominant role of tobacco in HNC development, risk differences across subsites, and potential interactions with other risk factors, HNC offers a particularly interesting opportunity to investigate the effects of tobacco exposure. In this context, the analysis of mutational signatures is an effective tool to track the complex mutagenic patterns linked to this and other exposures over a patient’s lifetime^11–13^. Certain mutational signatures have been related to well-established biological mechanisms and exposures. Signatures SBS4, found predominantly in lung cancer, and SBS92, in bladder cancer, capture two distinct mutagenic processes linked to tobacco use^12,14,15^. Conversely, Signature SBS16 has been attributed to alcohol consumption in esophageal and liver cancer^13,16^.

Previous studies exploring the genomic landscape of HNC have relied predominantly on exome sequencing data, which has limited power to detect mutational signatures, lacked a diverse geographical and ethnic representation of cases, and/or were limited to specific anatomical subsites^17–20^. Therefore, the carcinogenic mechanisms underpinning this cancer type in different geographical regions and anatomical subsites remain unclear. To bridge this gap, we performed whole-genome sequencing of 265 HNC cases from individuals exposed to known and suspected risk factors across eight countries with varying incidence rates. By leveraging mutational signature analysis combined with extensive epidemiological data, we shed light on the complexity of tobacco-induced mutagenesis and its interplay with alcohol consumption and other HNC risk factors.

## RESULTS

### Case-series overview and multi-country study design

A total of 265 HNC cases were included in the study, comprising retrospective collections from eight countries in Europe and South America^6,21^ (**Figure 1**; **Supplementary Table 1**). These encompass a broad geographic representation of HNC, including cases from high-incidence regions, with sex-combined age-standardized rates (ASR) ranging from 9.4 per 100,000 to 18.2 per 100,000 in Romania, Slovakia, Czech Republic, and Brazil, as well as moderate-incidence regions, with ASR from 3.8 to 7.8 per 100,000, in Colombia, Argentina, Greece, and Italy^1^. The dataset contains cases from all HNC anatomical subsites, with 127 oral cavity, 46 oropharynx, 17 hypopharynx, and 75 larynx cancers. Epidemiological questionnaire data were available on exposure to known and suspected HNC risk factors, including cases from drinkers, smokers, with both exposures, and non-exposed. DNAs from paired tumor and blood samples were extracted and whole-genome sequenced to average coverage of 55-fold and 27-fold, respectively.

**Figure 1.**
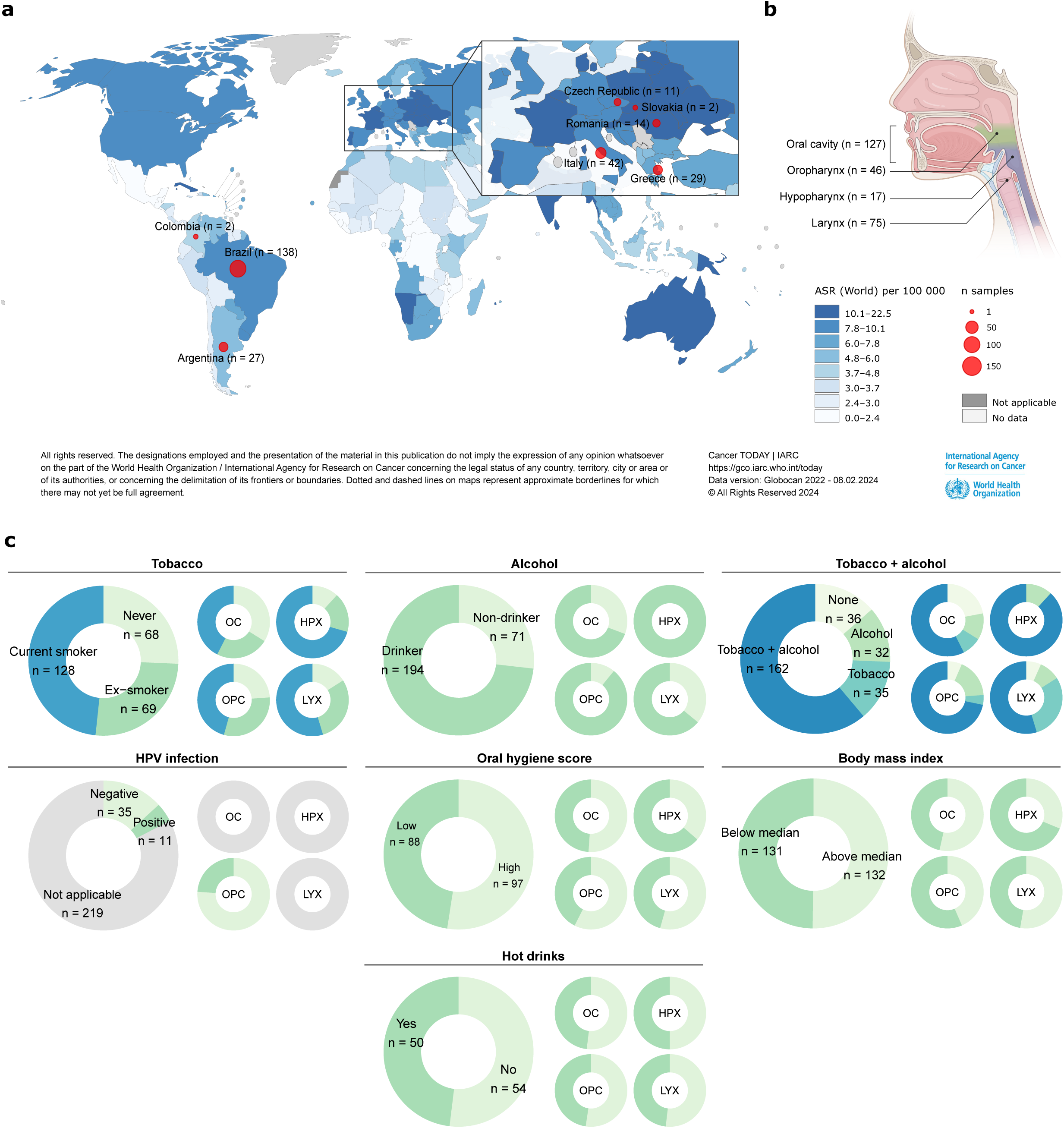
Head and neck cancer incidence and epidemiological characteristics. **a**, Incidence of head and neck cancer (HNC), sex-combined, age-standardized rates (ASR) per 100,000, data from GLOBOCAN 2022. Dots indicate countries included in this study and number of participating patients. **b,** Anatomical subsites of HNC, with number of tumor samples indicated in brackets. Created with biorender.com **c**, Known and suspected risk factors included in the study, based on epidemiological questionnaire data and human papillomavirus (HPV) detection. Frequencies of risk factors in the complete dataset (left) and by anatomical subsite (right) are indicated. OC, oral cavity; OPC, oropharynx; HPX, hypopharynx; LYX, larynx.

### Mutation burden

Among the 265 HNC cases, we observed a median of 12,887 single-base substitutions (SBS, range: 720 to 244,026), 63 doublet-base substitutions (DBS, range: two to 7,113), and 757 small insertions and deletions (ID, range: 124 to 9,898; **Supplementary Table 2**). Tumor samples from tobacco users exhibited higher SBS, DBS, and ID burdens compared to non-smokers (**Extended Data Figure 1b**; **Supplementary Table 3**), as previously reported for larynx cancer^14^. Differences were also found between anatomical subsites, with larynx samples presenting higher mutation burdens, even after correcting for tobacco status (**Extended Data Figure 1a**; **Supplementary Table 3**). No significant differences were found between geographical regions (**Extended Data Figure 1c**).

### Mutational signatures of exogenous and endogenous exposures

To investigate the mutational processes and carcinogenic exposures that have been operative in HNC development, we extracted SBS, DBS, and ID signatures and estimated the contribution of each signature to every sample. We obtained 15 *de novo* SBS signatures, which were decomposed into 18 reference signatures from the Catalogue of Somatic Mutations in Cancer (COSMICv3.2) database, and two signatures that could not be decomposed into any combination of existing signatures, SBS_I and SBS_L (**Figure 2a-b**; **Extended Data Figure 2**; **Supplementary Tables 4**,**7**-**9**, **Supplementary Note**).

**Figure 2.**
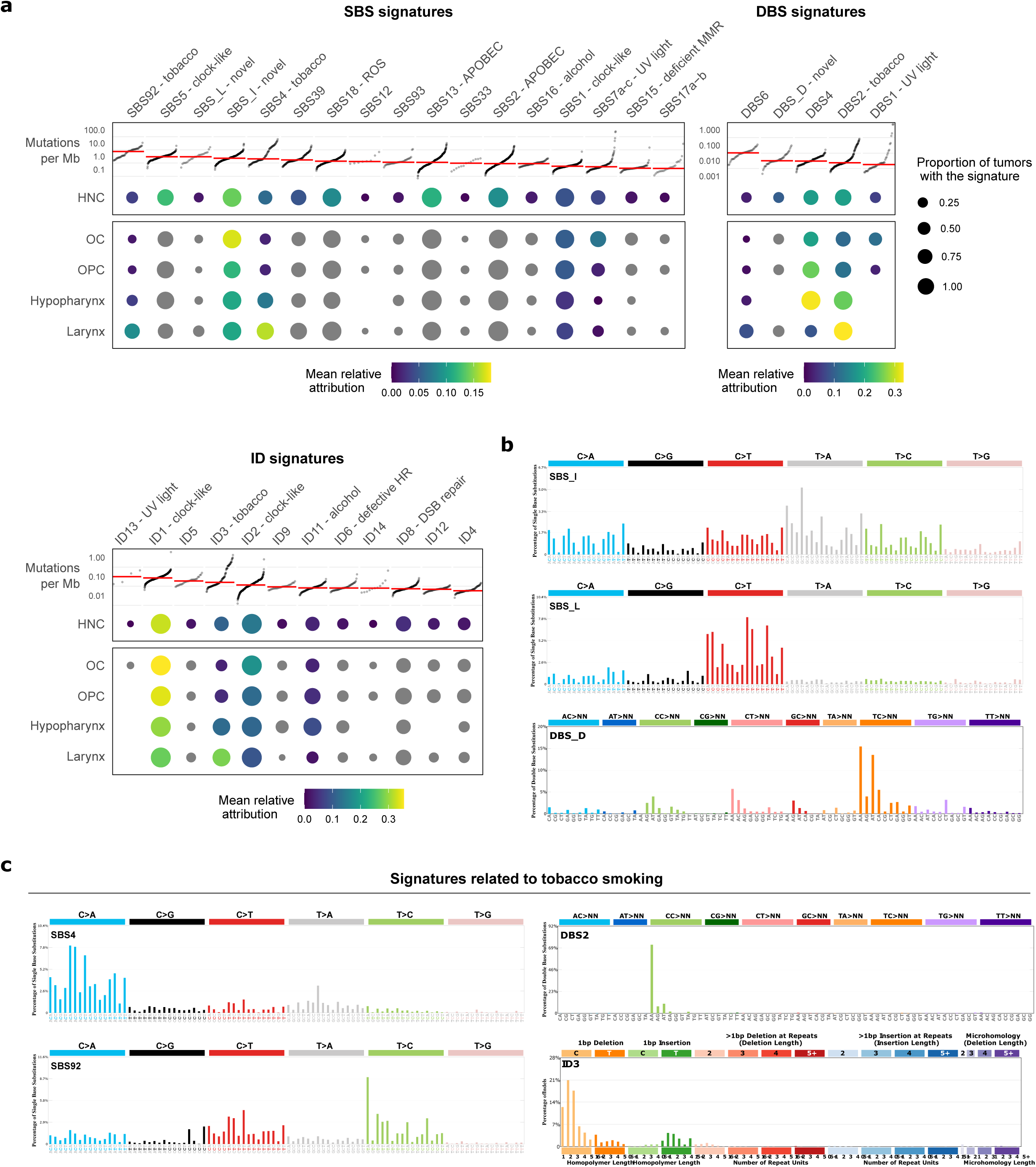
Mutational signature landscape of head and neck cancer. **a**, Single base substitution (SBS), doublet base substitution (DBS), and insertion deletion (ID) signatures extracted in 265 HNC tumors. The size of each dot represents the proportion of samples presenting each mutational signature in the whole HNC dataset and across anatomical subsites. The color represents the mean relative attribution of each signature. Gray dots indicate signatures without significantly different relative burdens by subsite. The top panel shows the mutations per megabase attributed to each signature in samples with counts higher than zero. Significance was assessed using a two-sided Kruskal-Wallis test and Bonferroni correction. **b**, Mutational spectrum of undecomposed signatures extracted from HNC. **c**, Known signatures of tobacco exposure identified in the HNC dataset. HNC, head and neck cancer; OC, oral cavity; OPC, oropharynx; ROS, reactive oxygen species; HR, homologous recombination; DSB, double-strand break.

Among the identified signatures, several have been previously associated with exogenous mutational processes^12^. The tobacco-related signatures SBS4 and SBS92 were found in 34% and 7.6% of HNC samples and respectively accounted for 6.3% and 3.5% of the mutational burden on average. SBS16, attributed to alcohol consumption^13,16^, was present in 19% of the samples with a modest impact on the HNC mutation burden of 1.4% on average. Signatures of ultraviolet (UV) light exposure SBS7a and SBS7b co-occurred in 4.2% of cases.

We also identified signatures associated with endogenous exposures and aberrant cellular processes. Notably, SBS2 and SBS13, which result from cytosine deamination by Apolipoprotein B mRNA-editing enzyme catalytic polypeptide-like (APOBEC)^14^, were present in the majority of HNC cases (93% and 92%, respectively; **Figure 2a**) and were highly correlated (**Supplementary Figure 1**). Combined, these signatures accounted for an average of 20.4% of the total SBS mutation burden. Other prevalent signatures included SBS18, which is caused by reactive oxygen species (77% of samples), and clock-like signatures SBS1 (78%) and SBS5 (55%) (**Figure 2a**).

Extraction of DBS signatures identified four *de novo* signatures, which decomposed into four COSMIC reference signatures (DBS1, DBS2, DBS4, and DBS6) and one non-decomposed signature (DBS_D; **Figure 2a-b**; **Extended Data Figure 3a**; **Supplementary Tables 5**,**7**-**9**). We also extracted seven *de novo* ID signatures, all of which were decomposed into 12 COSMIC signatures (**Figure 2a-b**; **Extended Data Figure 3b**; **Supplementary Tables 6**-**9**). DBS and ID signatures of exogenous exposures were positively correlated with their SBS counterparts (**Supplementary Figure 1**). For instance, the known tobacco-related signatures DBS2 (59% of samples) and ID3 (41%), along with DBS6 which has been previously registered as of unknown etiology, correlated with both SBS4 and SBS92. These associations are consistent with these SBS, DBS, and ID signatures being generated by the same underlying mutational process. Similarly, ID11 (38%), which was associated with alcohol consumption in esophageal cancer^13^, exhibited a positive correlation with the alcohol signature SBS16, while UV-related DBS1 (16.6%) and ID13 (1.5%) signatures showed the same link with SBS7a-c.

To establish which mutagenic exposures were active earlier or later during the development of HNC, we estimated the molecular timing of each SBS signature (**Methods**). Signatures of tobacco and alcohol consumption, as well as the SBS_L signature, were enriched in early clonal mutations (**Extended data Figure 4**), consistent with carcinogenic exposures occurring in normal cells^22^. Similarly, SBS_I was significantly enriched in early clonal mutations in oral cavity cases, while no significant differences were seen in other subsites. Signatures of APOBEC signaling and SBS39 were enriched in late clonal mutations, suggesting that the corresponding mutational processes increased in activity during the evolution of cancer clones^22^.

### HNC tumors present complex tobacco-related mutation patterns

Several signatures were independently associated with tobacco consumption, including the previously-recognized tobacco-related signatures SBS4, SBS92, DBS2, and ID3, as well as signature DBS6, reported as of unknown etiology, and the newly-discovered SBS_I (**Figure 3a-b**; **Supplementary Table 10; Supplementary Note**). The tobacco-associated SBS signatures were composed of three different substitution patterns (predominantly C>A for SBS4, T>C for SBS92, and T>A for SBS_I; **Figure 2c**) and exhibited transcriptional strand bias (**Supplementary Figure 4**)^15,23^. This strand bias towards the transcribed strand often occurs as a result of transcription-coupled DNA repair and is found in mutations due to bulky adducts, caused by exogenous exposures such as tobacco smoke carcinogens^23^. Assuming this mechanism is responsible for the strand bias in SBS_I, this is indicative of adduct formation on adenine bases.

**Figure 3.**
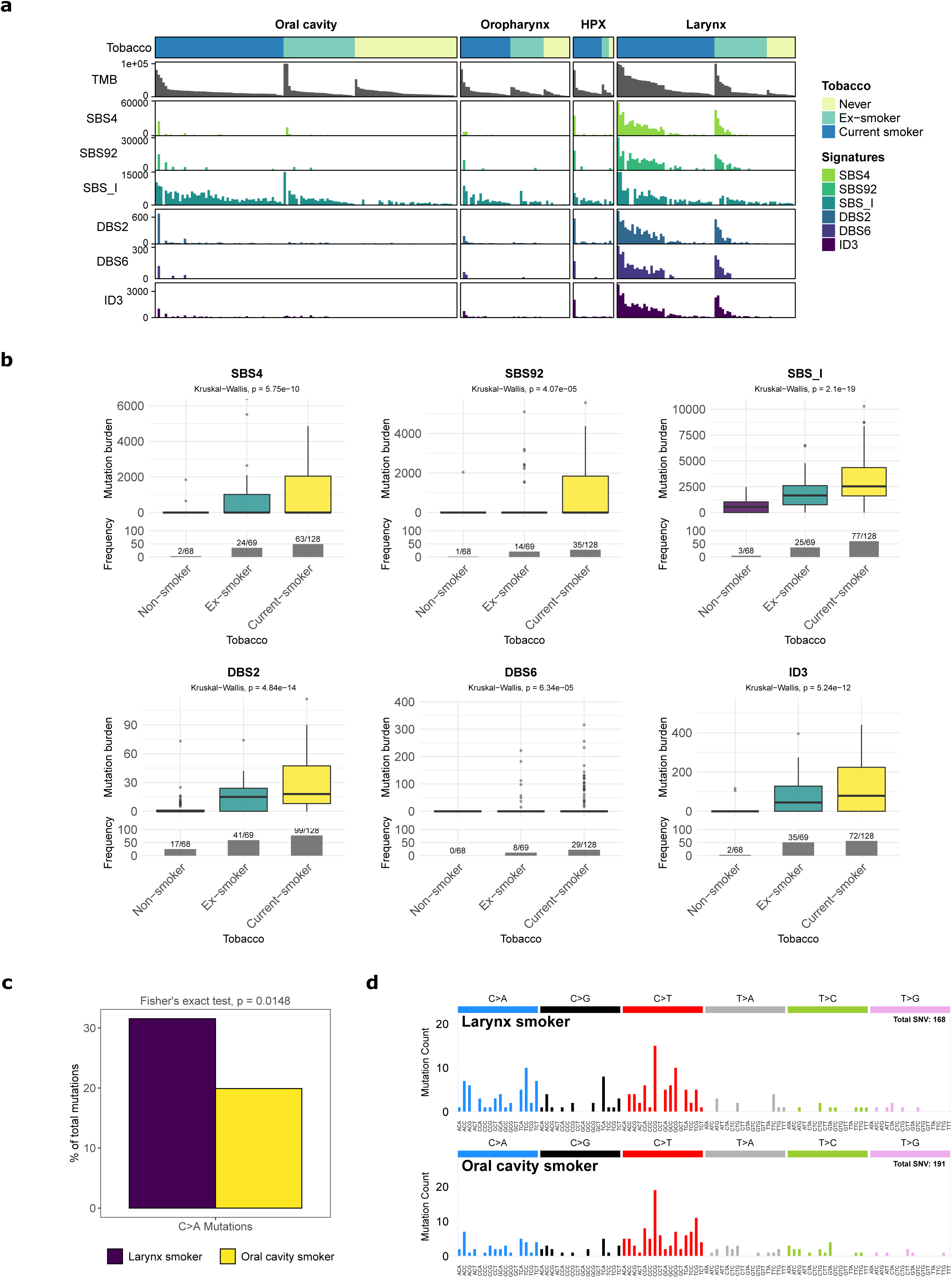
Tobacco-related signatures. **a**, Mutational burdens of tobacco-related signatures in HNC cases sorted by subsite and tobacco status. Tumor mutational burdens (TMB) per sample is also displayed. For clarity, y axis has been cut at 100,000 for TMB and at 15,000 for SBS_I**. b,** Mutational burdens for SBS, DBS, and ID signatures showing significant positive associations with tobacco consumption (n = 265 biologically independent samples). The Kruskal–Wallis test (two sided) was used to test for global differences. Box-and-whisker plots are in the style of Tukey. The line within the box is plotted at the median, while upper and lower ends indicate 25^th^ and 75^th^ percentiles. Whiskers show 1.5 × interquartile range (IQR), and values outside it are shown as individual data points. Y axes were cut at 1.25 × upper whisker for clarity. Bar plots indicate the frequencies of dichotomized signatures. **c,** Percentage of driver mutations occurring in C>A contexts in larynx and oral cavity HNC from smokers. **d,** SBS96-mutation spectrum of driver mutations in larynx and oral cavity HNC from smokers, showing enrichment in the frequency of C>A driver mutations in larynx cases. HPX, hypopharynx.

The distribution of tobacco-associated signatures varied across different anatomical subsites (**Figure 3a**, **Extended Data Figure 5a**; **Supplementary Table 10**). Previously established tobacco signatures exhibited higher signature burdens and frequencies in larynx cases compared to other subsites. For instance, SBS4 was present in 17% of OC, 17% of oropharynx, 53% of hypopharynx, and 67% of larynx cases. Similar distributions were observed for SBS92, DBS2, and ID3. Conversely, the previously unknown SBS_I signature was present in smokers across all subsites, with particular enrichment in the oral cavity. The associations between signatures and subsites remained significant after correction for tobacco consumption and other confounding variables **(Supplementary Table 10**).

### Effects of tobacco exposure on the driver mutation spectra

We explored the driver mutation profile in tobacco-related HNC. This revealed 96 cancer genes with driver mutations in our dataset, including *TP53, NOTCH1, CDKN2A, KMT2D*, and *CASP8*, which are commonly implicated in HNC^24^ (**Extended Data Figure 6a-b**; **Supplementary Tables 11-12**). *TP53* mutations were significantly enriched among smokers compared to non-smokers (83% [164/197] vs 61% [42/68], p=0.001), while *CASP8* mutations were more frequent among non-smokers (6.09% [12/197] vs 20.6% [14/68], p=0.003). A total of 642 driver mutations were identified (**Methods**), and these showed an enrichment of C>A substitutions in smokers compared to non-smokers (24.9% [114/457] vs 17.3% [32/185], Fisher’s exact test p=0.0379; **Extended Data Figure 6c**), consistent with the SBS4 mutation profile^12^. The frequency of C>A driver mutations in tobacco-exposed cases was higher in the larynx subsite compared to oral cavity (31.5% [53/168] vs 19.9% [38/191], Fisher’s exact test p=0.0148; **Figure 3c-d**). This reflects the lower contribution of SBS4 to mutations in tobacco-exposed oral cavity HNC compared to larynx cases, which has been carried through into the generation of driver mutations. T>A driver mutations were also observed among smokers, albeit in low frequencies (6.6% [11/168] in larynx and 8.4% [16/191] in oral cavity, hinting at a lower presence of SBS_I in driver mutations.

### Tobacco-related mutational signatures correlate with demographic HNC incidence

We analyzed the link between tobacco mutagenesis and variations in HNC incidence across different countries, sexes, and anatomical subtypes. Our findings support previous epidemiological evidence, which has shown a connection between HNC incidence and smoking habits^2^ (**Figure 4a-b**). Moreover, HNC incidence correlated with tobacco-related signatures (**Figure 4c**; **Supplementary Figure 2**), showing a higher ASR of HNC incidence in demographic groups presenting higher signature burdens. This further confirms that the geographical and demographic differences in tobacco exposure play a dominant role in driving HNC incidence.

**Figure 4.**
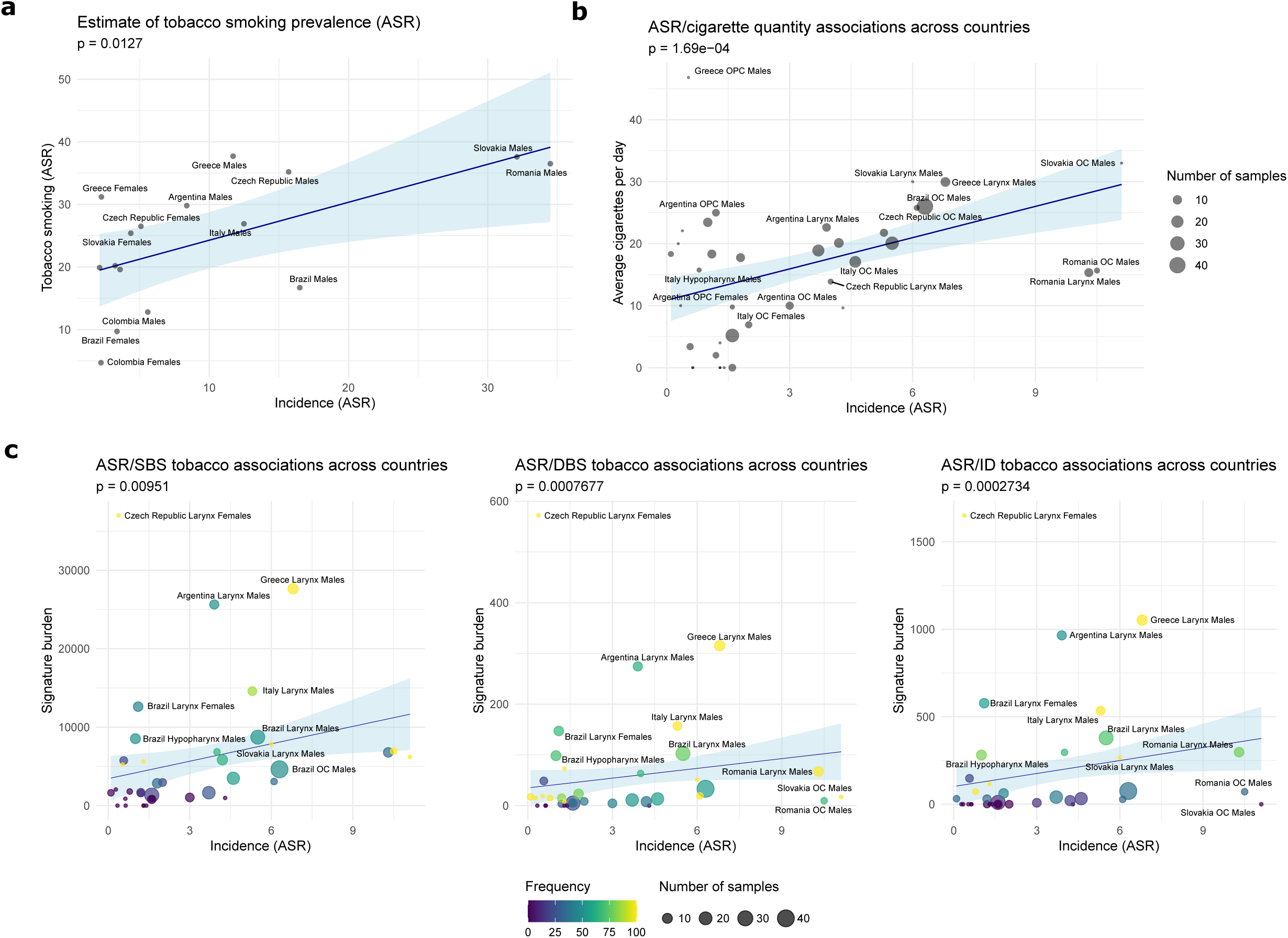
Association of tobacco use with incidence of head and neck cancer. **a**, Correlation between age-standardized rate (ASR) of HNC incidence and tobacco smoking per country and sex. Estimate of ASR of tobacco smoking prevalence was obtained from WHO Global Health Observatory (2019). **b**, Association between cigarette quantity smoked per day in the HNC dataset and ASR incidence per country, sex, and subsite. **c**, Association of tobacco-related signatures with ASR incidence. Number of mutations attributed to tobacco-related SBS (SBS4, SBS92, SBS_I), DBS (DBS2, DBS6), and ID (ID3) mutational signatures against ASR of HNC per country, sex, and subsite. For b and c, The p-values shown are for ASR variable in regressions across all cases, adjusted for age. The frequency of the signatures and number of cases per country, sex, and subsite are indicated. OC, oral cavity; OPC, oropharynx.

### Alcohol-related mutational signatures in drinkers and smokers

Next, we assessed the signature profile in HNC cases with a history of alcohol intake. Our analysis revealed significant associations between alcohol consumption and three specific signatures: SBS16, ID11, and DBS4 (**Figure 5**; **Supplementary Table 10**; **Supplementary Note**). SBS16 was present exclusively in HNC cases from drinkers and showed enrichment in samples exposed to both tobacco and alcohol compared to alcohol alone (29.0% [47/162] and 12.5% [4/32], respectively). Similarly, DBS4 and ID11 also presented higher burdens and signature frequencies in cases exposed to both risk factors (**Figure 5**; **Supplementary Table 10**). Although the etiology of DBS4 is unclear, it has been found prevalent in esophageal cancer cases from countries with high alcohol intake rates^13^. The results are, therefore, consistent with SBS16, DBS4 and ID16 all being generated by the same underlying alcohol-related mutational process and that the mutagenicity of this process is increased with co-exposure to tobacco smoke.

**Figure 5.**
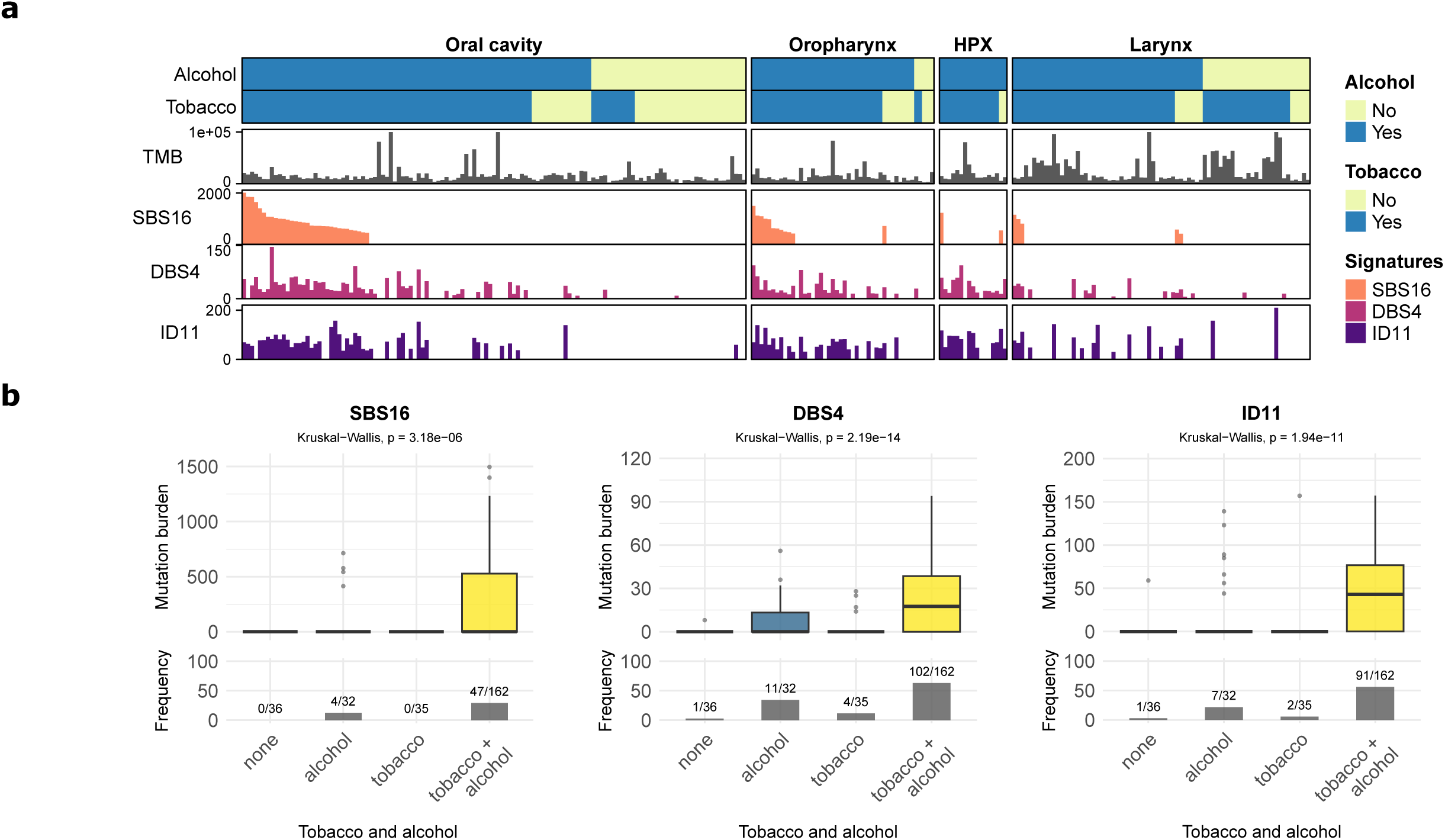
Alcohol-related signatures. **a**, Mutational burdens of tobacco-related signatures in HNC cases sorted by subsite, alcohol, and tobacco status. Tumor mutational burdens (TMB) per sample is also displayed. For clarity, y axis has been cut at 100,000 for TMB**. b,** Mutational burdens for SBS, DBS, and ID signatures showing positive associations with the tobacco plus alcohol status (n = 265 biologically independent samples). The Kruskal–Wallis test (two sided) was used to test for global differences. Box-and-whisker plots are in the style of Tukey. The line within the box is plotted at the median, while upper and lower ends indicate 25^th^ and 75^th^ percentiles. Whiskers show 1.5 × interquartile range (IQR), and values outside it are shown as individual data points. Y axes were cut at 1.25 × upper whisker for clarity. Bar plots indicate the frequencies of dichotomized signatures.

For driver mutations, samples from individuals exposed to both tobacco and alcohol were characterized by a particularly high *TP53* frequency of mutations (87.0% [141/162], 71.4% [25/35], 68.8% [22/32], and 55.6% [20/36] in the tobacco plus alcohol, alcohol alone, tobacco alone, and unexposed groups, respectively, Fisher’s exact test p=0.0001; **Extended Data Figure 6; Supplementary Table 11**). The driver mutation burden in the SBS16 context was too low to assess differences in the driver spectra between groups. However, *TP53* mutations in the SBS16 contexts were exclusively found in samples from individuals exposed to both tobacco and alcohol (*n*=5 *TP53* variants).

### HPV-positive HNC is characterized by APOBEC signatures

HPV infection in oropharynx cases did not elicit a specific mutational signature profile (**Supplementary Table 10**). However, the substitution profile in HPV-positive oropharyngeal HNC was characterized by a higher relative proportion of APOBEC signatures, with 57.6% of the signature burden being attributed to SBS2 and SBS13 on average, compared to 30.0% in HPV-negative oropharynx (**Extended Data Figure 7a**) consistent with previous reports^18^. Notably, the presence of APOBEC signatures was nearly ubiquitous across HNC cases (**Figure 2a**), suggesting a broader role for APOBEC activation beyond its anti-viral function^11^.

We also observed differences between HPV-positive and HPV-negative oropharynx cases exposed to tobacco. Among smokers, only 1/6 (17%) HPV-positive oropharynx cases presented tobacco-related SBS signatures, compared to 7/26 (27%) in HPV-negative cases (Fisher’s exact test p=0.0214). Despite the well-known influence of tobacco smoking on the driver profiles of HNC^20,24^, the driver alterations in HPV-positive smokers differed from that of HPV-negative smokers, and instead resembled the profile in HPV-positive cases from non-smokers. This included *PIK3CA* mutations, *PTEN* mutations and deletions, as well as absence of *TP53* mutations and of *FADD* gains (**Extended Data Figure 7b-c**). This, together with the reduced presence of tobacco-related signatures, suggests that oncogenesis in HPV-positive smokers may primarily be driven by viral infection rather than tobacco exposure.

### Mutational signature profile in samples exposed to putative HNC risk factors

We next investigated the presence of additional environmental exposures beyond the most widely-known HNC risk factors. Notably, UV-related signatures SBS7a-c, DBS1, and ID13 were detected predominantly in oral cavity cases (**Figure 6a**; **Supplementary Table 10**; **Supplementary Note**). SBS7 signatures have been previously described in HNC, but the anatomical and epidemiological features of positive cases have not been previously investigated^12^. Samples with a relative SBS7a-c burden of >10% were categorized as positive for UV exposure, a criterion met by 13 oral cavity cases from the lip, tongue, and floor of the mouth (**Figure 6b**). All positive cases were either tobacco or alcohol users, with 11/13 presenting both risk factors (**Figure 6b**). Thus, our data suggests a potential role of UV light exposure in HNC carcinogenesis^23^, which could be enhanced by tobacco and/or alcohol.

**Figure 6.**
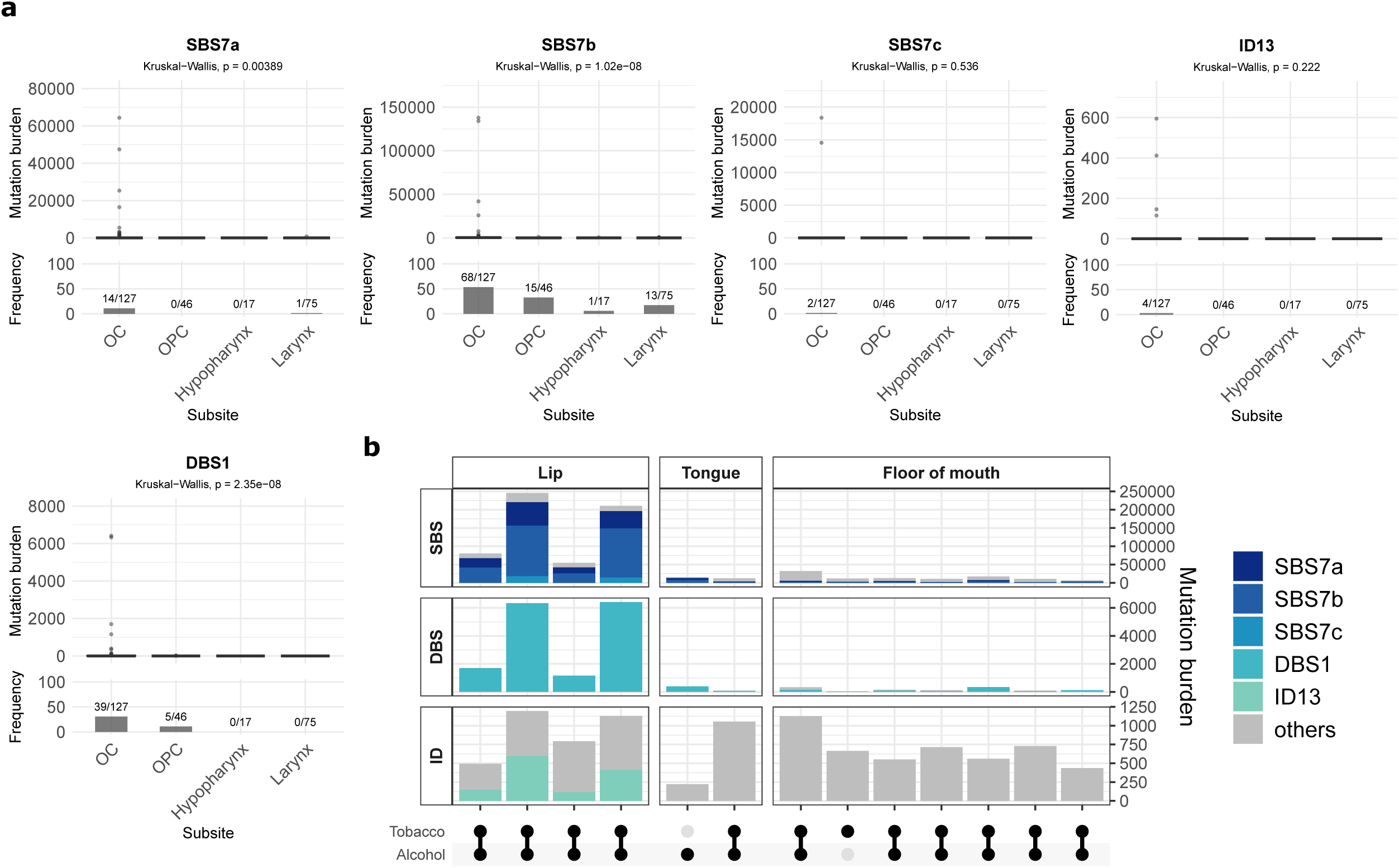
UV-related signatures in head and neck cancer. **a**, Mutational burdens for mutational signatures related to UV light exposure showing positive associations with the HNC anatomical subsite (n = 265 biologically independent samples). The Kruskal–Wallis test (two sided) was used to test for global differences. Box-and-whisker plots are in the style of Tukey. The line within the box is plotted at the median, while upper and lower ends indicate 25^th^ and 75^th^ percentiles. Whiskers show 1.5 × interquartile range (IQR), and values outside it are shown as individual data points. Frequencies of positive samples in each category are indicated in bar plots. **b,** Single base substitutions (SBS), doublet base substitutions (DBS) and small insertions and deletions (ID) signature burdens in samples positive for UV exposure based on relative SBS7a-c contributions above 10% of relative mutational burdens. Samples are sorted by lip (inner (*n*=3) or unspecified (*n*=1)), tongue, and floor of the mouth location within the oral cavity. Positive tobacco and alcohol status are indicated in black. OC, oral cavity; OPC, oropharynx.

Our analysis did not show any specific mutational patterns associated with other putative HNC risk factors, including hot drink consumption, poor oral health score, and high body mass index^6,7^ (**Supplementary Table 10**). This suggests that these agents are likely not causing direct mutagenesis. Finally, the previously unknown DBS_D signature and ID4, with unknown etiology, were enriched among non-smokers (**Extended Data Figure 5b**), suggesting a potential link to unidentified mutational processes in this population.

### HNC risk factors elicit distinct copy number profiles

HNC is characterized by complex patterns of copy number (CN) aberrations throughout the genome^19,20^. Unsupervised hierarchical clustering analysis on the CN counts in HNC samples (*n*=242) revealed two main clusters, one displaying diploid genomes (cluster D), and another presenting polyploidy and high burden of CN gains and losses (cluster P; **Extended Data Figure 8; Supplementary Figure 3**). These clusters further subdivided into four groups (D1, D2, P1, and P2). Notably, subgroup D2 was characterized by a CN-neutral profile, exhibiting significantly lower burdens of CN events compared to the other groups.

The CN clusters were associated with distinct epidemiological profiles (**Figure 7c-d**; **Supplementary Table 13**). Specifically, tobacco-related HNC were enriched within both the diploid and polyploid CN-high clusters (i.e., D1, P1, and P2), while the CN-silent cluster D2 was mostly constituted by samples from non-smokers, including cases with unknown risk factors and alcohol drinkers in the absence of tobacco. Consistent with this pattern, the D2 cluster was enriched in samples from female patients, oral cavity cases, and older age, aligning with the characteristic features of HNC with undefined risk factor^24^. Finally, HPV-positive oropharynx cases were enriched in the diploid clusters, predominantly in cluster D1.

**Figure 7.**
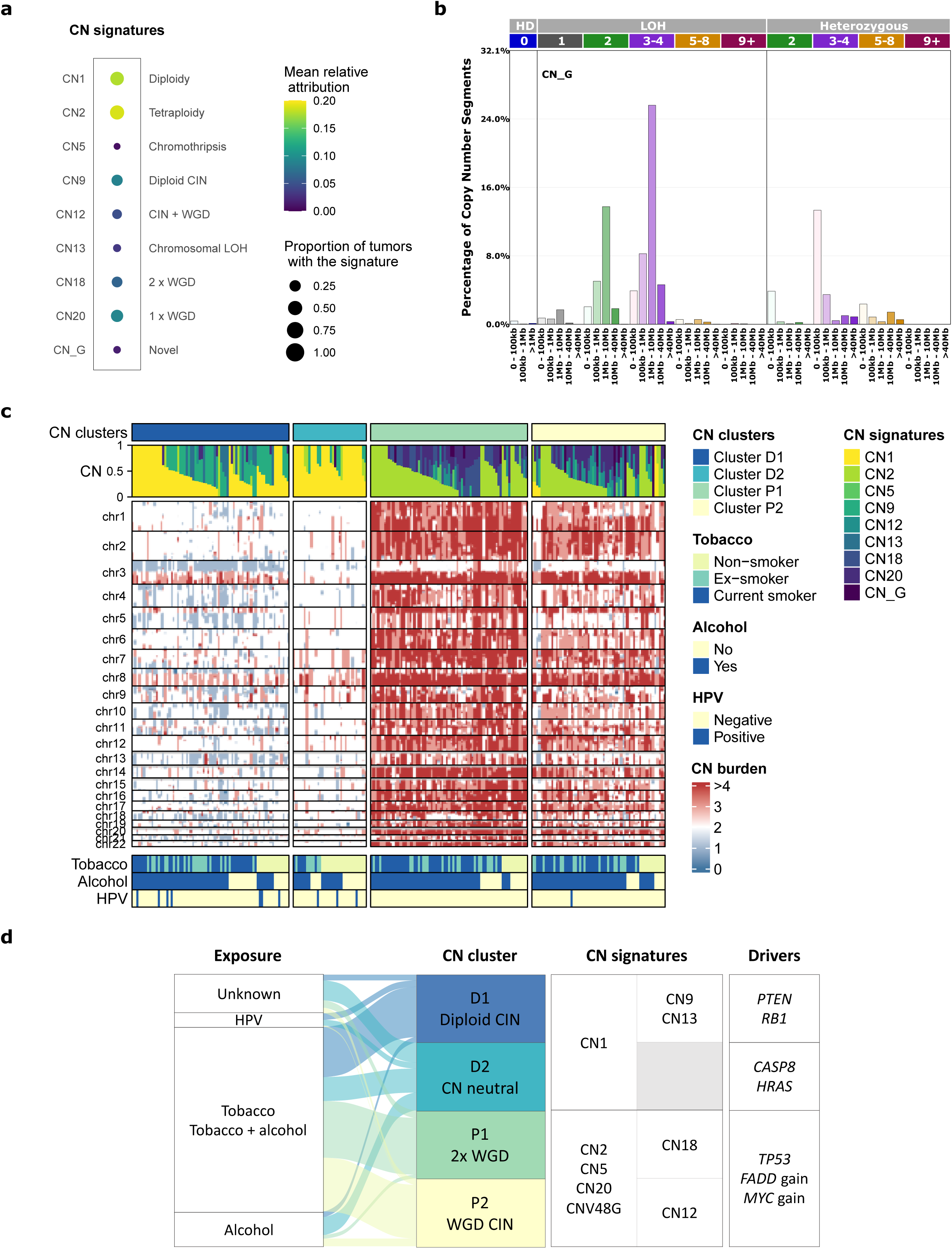
Copy number profile and copy number signature analysis in head and neck cancer. **a**, Copy number (CN) signatures extracted in 242 HNC tumors. The size of each dot represents the proportion of samples presenting the signature, and the color represents the mean relative attribution of each signature. **b,** Copy number spectrum of the newly-identified signature CN_G, defined by a 48 context copy number classification incorporating loss-of-heterozygosity status, total copy number state, and segment length to categorize segments from allele-specific copy number profiles. **c,** Copy number profiles of HNC cases classified by copy number cluster. Relative signature burdens, CN burden and associated epidemiological characteristics are indicated. The displayed epidemiological variables show significant differences by CN cluster as per Fisher’s exact test and Benjamini-Hochberg procedure. **d,** Summary of exposures, driver alterations and CN signatures associated with each CN cluster. Alluvial diagram depicts the frequency of each etiology in the CN clusters. WGD, whole-genome duplication; CIN, chromosomal instability; LOH, loss of heterozygosity.

To unveil distinct CN particularities within each CN cluster and etiology, we conducted CN signature analysis^25^ (**Figure *7*a-b; Extended Data Figure 9; Supplementary Note**). Cluster D1 exhibited enrichment in signatures of chromosomal instability within a diploid genome background (signatures CN1, CN9 and CN13; **Extended Data Figure 10a**). In contrast, cluster D2 presented a signature profile related to a diploid copy-neutral background (CN1). Clusters P1 and P2 displayed associations with signatures of whole-genome duplication (CN2, CN20) along with genomic aberrations (CN5, CN_G; **Extended Data Figure 10b-c**). Cluster P1 was consistent with double whole-genome duplications (CN18), while P2 showed signatures of chromosomal instability in conjunction with genome doubling (CN12). Collectively, our analysis suggests that HNC risk factors align with different CN profiles and provides an enhanced characterization of the CN aberrations in each HNC etiology (**Figure 7d**). Specifically, tobacco use, alone or with alcohol, may trigger chromosomal instability and aneuploidy, while HPV infection may confer a CN unstable diploid profile. Lastly, samples with unknown risk factors exhibit a CN-neutral profile.

We explored whether this difference in the CN profile could be due to the driver profile that is associated with each risk factor (**Figure 7d**; **Extended Data Figure 10d**). *TP53* mutations and *MYC* gains, two known promoters of genomic instability^25–27^, as well as gains in the anti-apoptotic *FADD* gene, were enriched in cluster P. *CASP8* and *HRAS* mutations were enriched in the D2 CN-neutral cluster, in agreement with previous studies in HNC^20,24,25^. Finally, *PTEN* and *RB1* mutations were enriched in the D1 cluster. Overall, these results show that tobacco use in HNC is associated with a distinct CN-rich profile and driver alterations related to genome instability.

## DISCUSSION

The role of tobacco as one of the most avoidable cancer risk factors has been known for over 50 years. Yet, the detailed mechanisms by which tobacco smoke leads to DNA damage and carcinogenesis in different tissues are still not fully understood^14,28,29^. In this study encompassing HNC cases from eight countries in Europe and South America, we shed light on the effects of tobacco as the main mutagenic exposure in HNC and explored the complex mutational patterns and genomic alterations linked to tobacco exposure in different HNC subsites, as well as its interplay with alcohol consumption and other risk factors.

Tobacco smoke contains a mixture of thousands of chemicals, including over 60 carcinogens, among which benzo[*a*]pyrenes (BaP) and nitrosamines are the most widely studied. These carcinogens undergo metabolic activation, generating reactive intermediates that interact with DNA in exposed tissues, resulting in complex mutagenic processes that can lead to cancer development^29^. In HNC, tobacco exposure resulted in six different signatures, unveiling at least three mutational processes due to tobacco in HNC. Signature SBS4, characterized by C>A transversions, has been largely attributed to BaP adducts^14,30,31^. Exposure to this compound is also consistent with the CC>AA substitutions and C deletions present in DBS2 and ID3 tobacco signatures, respectively^31^. Conversely, signature SBS92, composed predominantly of T>C transitions, has not been related to specific carcinogens in tobacco smoke^15^. Finally, the T>A-rich substitution profile captured by the previously-unidentified signature SBS_I is compatible with adduct formation on adenines, which have been observed in response to multiple tobacco compounds^31–33^. Among those, exposure to nicotine-derived nitrosamine ketone (NNK), one of the main tobacco carcinogens in oral tissues^34^, also yielded a T>A-rich signature *in vitro*^35,36^. Notably, a signature exhibiting high T>A frequencies and transcriptional strand bias has been described in normal lung epithelia from patients with a history of smoking^37^.

Our epidemiological analysis revealed that the mutational effects of tobacco vary among anatomical subsites. The canonical tobacco signatures SBS4 and SBS92 were found predominantly in larynx cases, along with the tobacco-related DBS and ID signatures. Conversely, SBS_I was extracted in HNC cases from all subsites, with a notable enrichment in oral cavity cases. Altogether, our observations hint at varying susceptibility, exposure level, or clearance of tobacco carcinogens across tissues, leading to different genotoxic effects. A possible explanation for these differences is the tissue-specific pattern of cytochrome P450 function. CYP1A1, the main BaP metabolizer, is primarily expressed in lung and larynx, whereas enzymes responsible for nitrosamine metabolism, such as CYP2E1, are predominant in the upper aerodigestive tract including the oral cavity^34,38–40^. These differences in the response to tobacco across tissues may partially explain the greater susceptibility to smoking found for larynx cancers compared to other anatomical subsites^10^. While tobacco use was associated with elevated mutation burdens and BaP-related driver mutations in larynx cancers, this was not observed in oral cavity cases, aligning with a reduced carcinogenic effect. Thus, additional carcinogenic processes may be necessary to aid in the development of oral cavity and oropharynx cancers, including alcohol and HPV infection^10^.

In this regard, we also identified mutagenic processes linked to alcohol exposure^13,16^, including signatures SBS16, ID11, and an unreported association with signature DBS4. In HNC, alcohol-related signatures were predominantly observed in patients reporting both alcohol and tobacco consumption, reflecting the synergistic effect between these two factors on disease risk^9,41^. Furthermore, a previous study suggested an enrichment of SBS16 in oropharynx cases from tobacco users^18^. Altogether, our findings indicate that tobacco could enhance the carcinogenic effects of alcohol through shared mutagenic processes. Experimental evidence suggests that salivary concentrations of acetaldehyde, the genotoxic byproduct of alcohol metabolism, are greatly increased by tobacco smoking^42^, which could result in enhanced alcohol-related mutagenesis in cases with combined exposure.

Our data show that tobacco use, alone or in conjunction with alcohol, is also associated with a distinct CN-rich profile, characterized by CN signatures of chromosomal instability, and resembling a previously described subset of CN-rich HNC^19^. These genomic profiles are likely due to driver alterations leading to genome instability such as *TP53* mutations, which are prevalent among smokers and drinkers^24^. Although high CN burdens have been reported in lung adenocarcinoma cases from smokers^14^, the link between this exposure and specific CN or driver profiles in HNC was previously unclear^43^. Cases with unknown etiology, on the other hand, exhibit few CN alterations, prevalence of *CASP8* and *HRAS* mutations, and wild-type *TP53*. A similar CN-neutral group of samples has been observed in HNC, with an unreported link with HNC etiology^19,44^.

Regarding the mutagenic potential of other investigated risk factors, HPV infection did not elicit a specific mutational signature profile, but it was associated with distinct driver mutations and a CN-unstable diploid genome. Poor oral hygiene, high body mass index, and consumption of hot drinks did not display a direct effect on the mutation profile of HNC cases and likely contribute to the development of HNC of unknown etiology through mechanisms distinct from direct mutagenesis. This pattern has been proposed for several carcinogens in prior studies^13,45^. Nevertheless, there may exist additional unidentified mutagens leading to HNC, as hinted by the presence of the previously unidentified signature SBS_L as well as the enrichment of DBS_D and ID4 among non-smokers.

Furthermore, we provide evidence suggesting that sunlight exposure may contribute to HNC development. Specifically, we identified signatures consistent with pyrimidine dimer formation (SBS7a-c and DBS1) in oral HNC cases, indicative of DNA damage by UV light^12,23^. UV light has only been described as a risk factor for malignancies in the external lip^46^, but experimental evidence suggests that oral cavity epithelia are susceptible to this exposure, and its carcinogenic processes could be enhanced by tobacco smoking^47–51^. While we cannot exclude the possibility of other mutational processes eliciting CC>TT substitutions, such as those driven by reactive oxygen species^52^, the presence of ID13 signatures, identified in melanoma^12^, provides additional evidence supporting the role of sunlight exposure in oral HNC.

In summary, through our comprehensive analysis of the mutational, genomic, and epidemiological profile of HNC cases from diverse geographical regions, we have uncovered genomic mechanisms by which tobacco smoke and other risk factors contribute to HNC development. These findings enhance our understanding of the complexity and tissue-specificity of tobacco mutagenesis, offering additional evidence that may inform prevention strategies aimed at reducing the risk of this disease.

## ONLINE METHODS

### Recruitment of cases and informed consent

The IARC/WHO coordinated participant recruitment through the HEADSpAcE and Central European international networks, comprising 13 collaborators from the eight participating countries in Europe and South America (**Supplementary Table 14**). Inclusion criteria for patients were ≥18 years of age (ranging from 18 to 90 years; with a mean of 60 and standard deviation of 12 years), confirmed diagnosis of primary HNC, and no prior cancer treatment. Informed consent was obtained for all participants. Patients were excluded if they had any condition that could interfere with their ability to provide informed consent or if there were no means of obtaining adequate tissues as per protocol requirements. Ethical approvals were first obtained from each local research ethics committee and federal ethics committee when applicable, as well as from the IARC Ethics Committee.

### Bio-samples and data collection

Dedicated standard operating procedures, following guidelines from the International Cancer Genome Consortium (ICGC), were designed by the IARC/WHO to select adequate retrospective case series with complete biological samples and exposure information as described previously^1,2^ (**Supplementary Table 14**). In brief, for all case series included, anthropometric measures were taken, together with relevant information regarding medical and familial history. All biological samples from retrospective cohorts were collected using rigorous, standardized protocols and fulfilled the required standards of sample collection defined by the IARC/WHO for sequencing and analysis. Retrospective case series were included after examination of their respective recruitment protocols to ensure the availability of necessary biological samples based on standard operating procedures, following guidelines from the ICGC, and also based on the collection of relevant exposure history based on a comparison of validated epidemiological questionnaires from each specific region. Comparable smoking and alcohol history was available from all centers, as well as detailed epidemiological information on oral health, coffee, tea, and mate consumption for specific regions^2^.

Potential limitations of using retrospective clinical data collected using different protocols from different populations were addressed by central data harmonization to ensure a comparable group of exposure variables (**Supplementary Table 15**). All patient-related data, as well as clinical, demographical, lifestyle, pathological, and outcome data, were pseudonymized locally using a dedicated alphanumerical identifier system before being transferred to the IARC/WHO central database.

### Expert pathology review

Original diagnostic pathology departments provided diagnostic histological details of contributing cases through standard abstract forms, together with a representative hematoxylin–eosin-stained slide of formalin-fixed paraffin-embedded tumor tissues whenever possible. The IARC/WHO centralized the entire pathology workflow and coordinated a centralized digital pathology examination of frozen tumor tissues collected for the study, as well as formalin-fixed paraffin-embedded sections when available, via a web-based report approach and a dedicated expert panel, following standardized procedures as described previously^1^. A minimum of 50% viable tumor cells was required for eligibility for whole-genome sequencing.

### DNA extraction

Extraction of DNA from fresh frozen tumor and matched blood samples was centrally conducted at IARC/WHO. Of the cases that proceeded to the final analysis (*n*=265), germline DNA was extracted from blood samples using previously described protocols and methods^1^.

### HPV infection status and genome detection

The HPV infection status was determined by HPV16 E1, E2, E6 and E7 serology. To assess the HPV status in oropharynx cases with missing serologic information (*n*=3), we used two orthogonal NGS-based viral integration tools: Virus intEgration sites through iterative Reference SEquence customization (VERSE) and Fast Viral Integration and Fusion Identification (FastViFi)^3,4^ (**Supplementary Table 16; Supplementary Note**). VERSE was utilized as part of the VirusFinder2.0 package: https://bioinfo.uth.edu/VirusFinder/ and FastViFi was installed using github: https://github.com/sara-javadzadeh/FastViFi. Default parameters were used for running both tools.

### Whole-genome sequencing

A total of 618 patients with HNC were enrolled in the study. Out of those, 315 cases were selected based on pathologic review and DNA quality (tumor and germline), and DNA was received at the Wellcome Sanger Institute for whole genome sequencing. To ensure the tumor and matched normal sample originated from the same individual, Fluidigm SNP genotyping with a custom panel was performed. Whole-genome sequencing (150 bp paired-end) was performed on the NovaSeq 6000 platform with target coverage of 40× for tumors and 20× for matched normal tissues. All sequencing reads were aligned to the GRCh38 human reference genome using Burrows-Wheeler-MEM (version 0.7.16a and version 0.7.17). A standard set of post-sequencing quality criteria was applied for metrics including total coverage, evenness of coverage, and contamination. Cases were excluded if coverage was below 30× for tumors or 15× for normal tissue. For evenness of coverage, the median over mean coverage (MoM) score was calculated, and tumor samples with MoM scores outside the range of values (0.92 – 1.09) which were determined by previous studies to be appropriate were excluded^5^. Conpair^5^ (https://github.com/nygenome/Conpair) was used to detect contamination, and any tumor or normal sample with a value above 3% was excluded^6^. A total of 265 cases passed all criteria and were included in subsequent analysis.

### Somatic variant calling

A standard analysis pipeline (https://github.com/cancerit) was used to perform variant calling for copy number variants (ASCAT^7^ and Battenberg^8^, when tumor purity allowed); SNVs (cgpCaVEMan^9^); Indels (cgpPindel^10^); and structural rearrangements (BRASS). CaVEMan and BRASS were run using the copy number profile and purity values determined from ASCAT when possible (complete pipeline, *n*=242) or using copy number defaults and an estimate of purity obtained from ASCAT– BATTENBERG when tumor purity was insufficient to determine an accurate copy number profile (partial pipeline, *n*=23). For SNVs, additional filters (ASRD ≥140 and CLPM = 0) in addition to the standard PASS filter. To further exclude the possibility of caller-specific artifacts being included in the analysis, a second variant caller, Strelka2, was run for SNVs and IDs^1,11^, with variants called by both the Sanger variant-calling pipeline and Strelka2 included in the final analysis.

### Generation of mutational matrices

Mutational matrices for single base substitutions (SBS), doublet base substitutions (DBS), small insertions and deletions (ID), and copy number variants (CNV) were generated using SigProfilerMatrixGenerator (https://github.com/AlexandrovLab/SigProfilerMatrixGenerator) with default options (v1.2.0)^12^.

### Mutational signature analysis

Multiple methods were used to extract mutational signatures. The primary extractions were performed using SigProfilerExtractor (https://github.com/AlexandrovLab/SigProfilerExtractor) with a second method mSigHdp used to validate the de novo mutational signatures extracted (https://github.com/steverozen/mSigHdp)^13,14^. SigProfilerExtractor v1.1.13 was run using nndsvd_min initialization (NMF_init="nndsvd_min") for 1-20 signature solutions and 500 NMF replicates. For SBS mutational signatures were extracted in both SBS1536 and SBS288 contexts. Both results were similar (**Supplementary Note**) with the SBS1536 results taken forward for the final analysis (**Supplementary Table 4**). Signatures were extracted using SigProfilerExtractor in the following contexts for other variant types; DBS78 for DBS, ID83 for indels, and CNV48 for copy number variants (**Supplementary Tables 5-6**,**17**). The extracted de novo signatures were decomposed to COSMIC reference signatures where possible; this step is important as it allows the detection of *de novo* signatures which are made up of multiple reference signatures that have not separated during the extraction process (**Supplementary Note**). mSigHdp extractions were performed using the suggested parameters and using the country of origin to construct the hierarchy for SBS96 and ID83 contexts. A comparison of the SigProfilerExtractor and mSigHdp results can be found in the **Supplementary Note**.

### Attribution of activities of mutational signatures

MSA v2.0 (https://gitlab.com/s.senkin/MSA) was used to attribute both de novo and COSMIC mutational signatures15. For COSMIC attributions the panel of signatures included reference signatures identified during the decomposition of mutational signatures in addition to newly extracted signatures which were not decomposed. A conservative approach was used for MSA attributions utilizing the (params.no_CI_for_penalties=false) option for calculation of optimum penalties. Pruned attributions were used for final analysis, where confidence intervals have been applied to each attributed mutational signature and any signature activity with a lower confidence limit equal to 0 are removed.

### Driver mutations

Driver mutations in HNC were identified using the following methods. Firstly, dNdS was used to identify genes under positive selection in HNC^16^. Results were calculated both for the whole genome (q-value<0.01), and with restricted hypothesis testing (RHT) for a panel of 369 known cancer genes^16^. Variants in any gene identified as under positive selection in global dNdS or in the 369-cancer gene panel were considered as potential driver mutations and were then classified as likely drivers if they met any of the following criteria: (i) Truncating mutations in genes annotated as tumor suppressors; (ii) mutations annotated as likely or known oncogenic in MutationMapper; (iii) truncating variants in genes with selection (q value<0.05) for truncating mutations assumed to be tumor suppressors and thus likely drivers; (iv) missense variants in all genes under positive selection and with dN/dS ratios for missense mutations above five (assuming four of every five missense mutations are drivers) labeled as likely drivers; or (v) in-frame indels in genes under significant positive selection for in-frame indels. The Cancer Gene Census (https://cancer.sanger.ac.uk/census) and the Cancer Genome Interpreter tool (https://www.cancergenomeinterpreter.org) were used to annotate potential drivers with the mode of action. Missense mutations were assessed using the MutationMapper tool (http://www.cbioportal.org/mutation_mapper).

### Copy number profile

The copy number profiles were investigated in a subset of cases with available copy number data (complete pipeline, *n*=242). Unsupervised clustering analysis of the copy number counts was performed using Euclidean distance and Ward’s agglomerative procedure. Driver copy number alterations were defined as cancer-related alterations in the COSMIC cancer gene census as follows^17,18^: (1) homozygous deletion (CN = (0, 0)) of genes listed as deleted in COSMIC; and (2) amplification (CN > 2 × ploidy + 1) of genes listed as amplified (A) in COSMIC or *PIK3CA* gains, a commonly-reported HNC alteration^19,20^.

### Evolutionary analysis

MutationTimeR^21^ was run to annotate mutations as either early clonal, late clonal, subclonal, or NA clonal (meaning clonality could not be assigned). Samples with at least 256 early clonal mutations and at least 256 late clonal mutations were retained (*n*=173), and the early and late clonal mutations for these samples were split into individual VCF files. SigProfilerAssignment^22^ was run on the resulting VCF files to identify the mutational processes active in the early clonal and late clonal mutations for each sample. Differences between the early and late relative activity of each mutational signature were assessed using a Wilcoxon signed-rank test, and p-values were corrected across signatures using the Benjamini-Hochberg Procedure (q-value).

### Regressions and associations with signatures

Signature attributions were dichotomized into presence and absence using confidence intervals, with presence defined as both lower and upper limits being positive, and absence as the lower limit being zero. If a signature was present in at least 75% of cases (SBS1, SBS2, SBS13, SBS18, SBS_I, ID1, and ID2), it was dichotomized into above and below the median of attributed mutation counts. The binary attributions served as dependent variables in logistic regressions, and relevant risk factors were used as factorized independent variables. Regressions with variables presenting data separation were performed using Firth’s penalized logistic regression.

For SBS, DBS, and ID mutation burden analyses, cases defined as hypermutators (mutation burdens more than 1.5 IQR above Q3) were excluded and associations with epidemiological factors were assessed using linear regression analysis.

Regressions with HNC incidence (age-standardized rates) were performed as linear regressions with signature attributions (those present in at least 75% of cases) with confidence intervals not consistent with zero. Signatures present in less than 75% of cases were dichotomized into presence and absence as previously mentioned and analyzed using the logistic regressions. Age-standardized rates were obtained from Global Cancer Observatory (GLOBOCAN 2022)^23^. Regressions were performed on a sample basis.

To adjust for confounding factors, sex, age of diagnosis, subsite, region, tobacco, and alcohol status were added as covariates in all regressions. The region variable was categorized as Europe and South America. The Bonferroni method was used to test for significant p-values.

## DATA AVAILABILITY

Whole genome sequencing data and patient metadata are deposited in the European Genome-phenome Archive (EGA) associated with study EGAS00001005450. Mutational catalogs for the PCAWG dataset can be accessed at https://dcc.icgc.org/releases/PCAWG. All other data is provided in the accompanying Supplementary Tables.

## CODE AVAILABILITY

All algorithms used for data analysis are publicly available with repositories noted within the respective method sections and in the accompanying reporting summary. Code used for regression analysis and figures is available at https://gitlab.com/mutographs-hnc.

## ACKNOWLEDGEMENTS

The authors would like to thank Laura O’Neill, Kirsty Roberts, Katie Smith, Siobhan Austin-Guest and the staff of Sequencing Operations at the Wellcome Sanger Institute for their contribution. We are grateful for the support provided by the IARC General Services, including the Laboratory Services and Biobank team led by Z. Kozlakidis, the Section of Support to Research overseen by T. Landesz. The authors would like to thank Maggie Blanks and Mimi McCord for useful discussions. The authors would also like to thank all the patients and their families involved in this study.

## CONTRIBUTIONS

The study was conceived and designed by S.P. and P.B., and supervised by S.P., P.B., M.R.S., and L.B.A. Analysis of data was performed by L.T., S.M., A.C.D.C., M.K., S.C., S.S., T.C., R.C.C.P., J.R.A., V.G., A.A., E.N.B., R.V., J.W., S.F., M.D., D.J., and J.T. Pathology review was carried out by B.A. Sample manipulation was carried out by P.C., C.Carreira, and C.L. Patient and sample recruitment was led or facilitated by F.R.P., L.P.K., J.P., F.G., J.C.D.C., P.L., A.L., M.V., D.M., I.N.M., L.M.A., R.R., J.R.V.P., S.V.V.Z., I.H., M.P.C., C.Canova, E.F., and P.A.R. Data generation was performed by J.W., S.F., and C.L. Scientific project management was carried out by A.C.D.C. and L.H. L.T. L.T. and S.M. contributed and were responsible for overall scientific coordination. The manuscript was written by L.T., S.M., A.C.D.C., L.B.A., P.B., M.R.S., and S.P., with contributions from all other authors.

## DISCLAIMER

Where authors are identified as personnel of the International Agency for Research on Cancer/ World Health Organization, the authors alone are responsible for the views expressed in this article and they do not necessarily represent the decisions, policy or views of the International Agency for Research on Cancer / World Health Organization.

## CORRESPONDING AUTHOR

Correspondence to Sandra Perdomo.

## EXTENDED DATA FIGURE LEGENDS

**Extended Data Figure 1.**
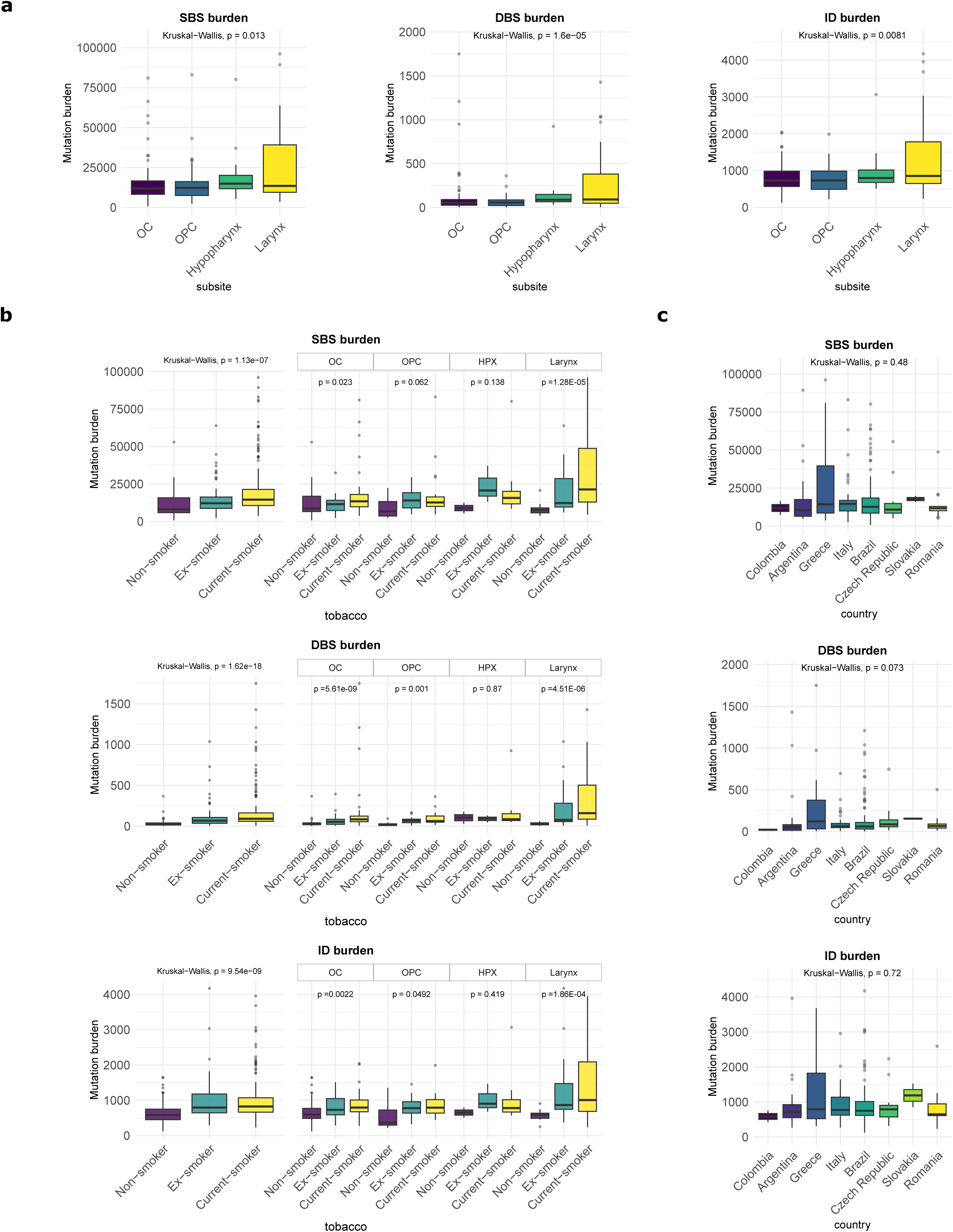
Mutational burdens in HNC. **a-c**, Mutational burdens for single base substitutions (SBS), doublet base substitutions (DBS) and small insertions and deletions (ID) burdens by anatomical subsite (a), smoking status (b) and country (c). Panel b depicts the mutation burdens by smoking status in the whole HNC dataset (left) and across anatomical subsites (right). Kruskal– Wallis test (two sided) was used to test for global differences. Box-and-whisker plots are in the style of Tukey. The line within the box is plotted at the median, while upper and lower ends indicate 25^th^ and 75^th^ percentiles. Whiskers show 1.5 × interquartile range (IQR), and values outside it are shown as individual data points. Hypermutators defined as samples with mutation burdens above 100,000 for SBS (*n*=4), 6,000 for DBS (*n*=1) and 5,000 for ID (*n*=1) were removed from the analysis. OC, oral cavity; OPC, oropharynx; HPX, hypopharynx.

**Extended Data Figure 2.**
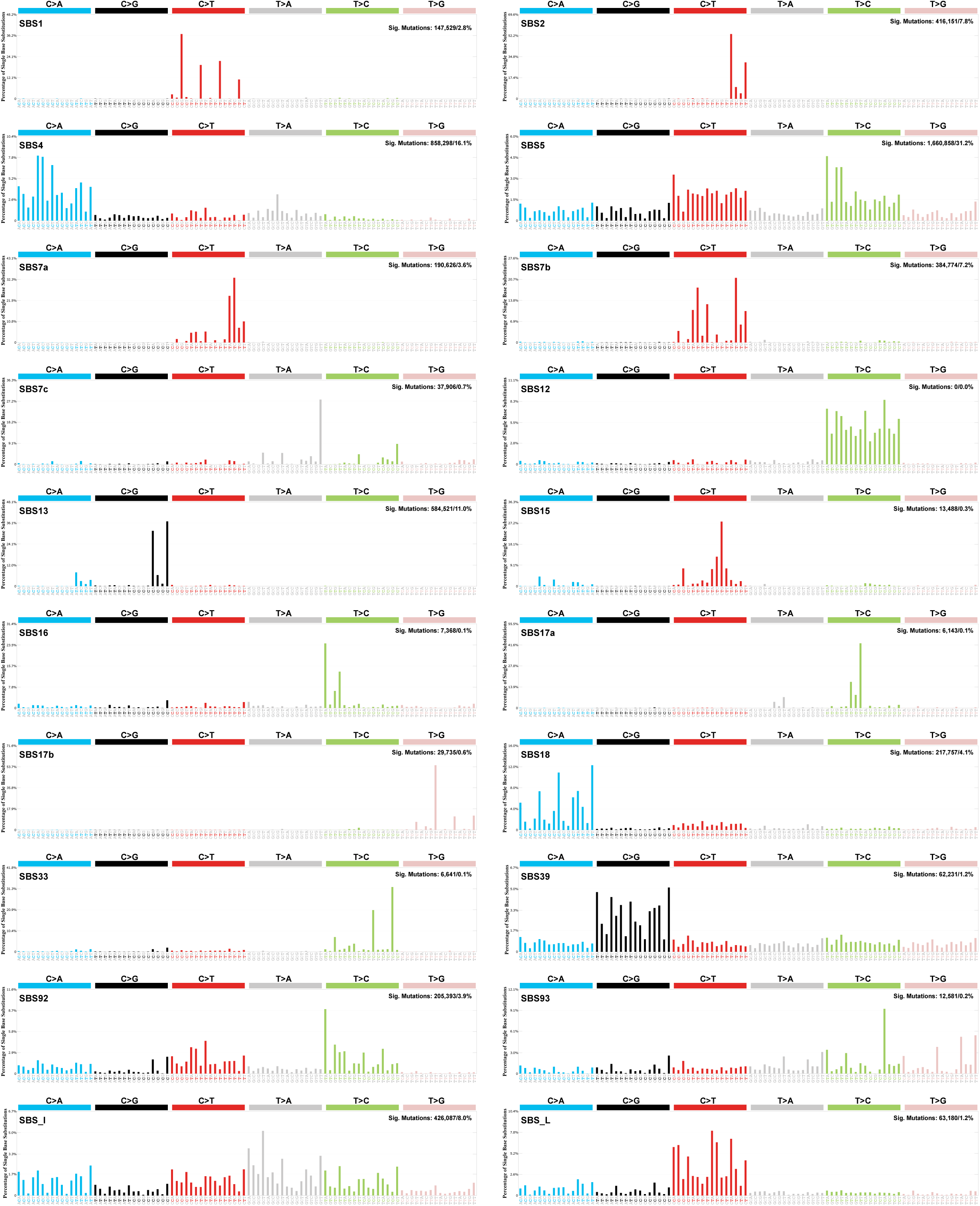
SBS signature decomposition. Decomposed SBS signatures, including reference COSMIC signatures and *de novo* signatures not decomposed into COSMIC reference signatures.

**Extended Data Figure 3.**
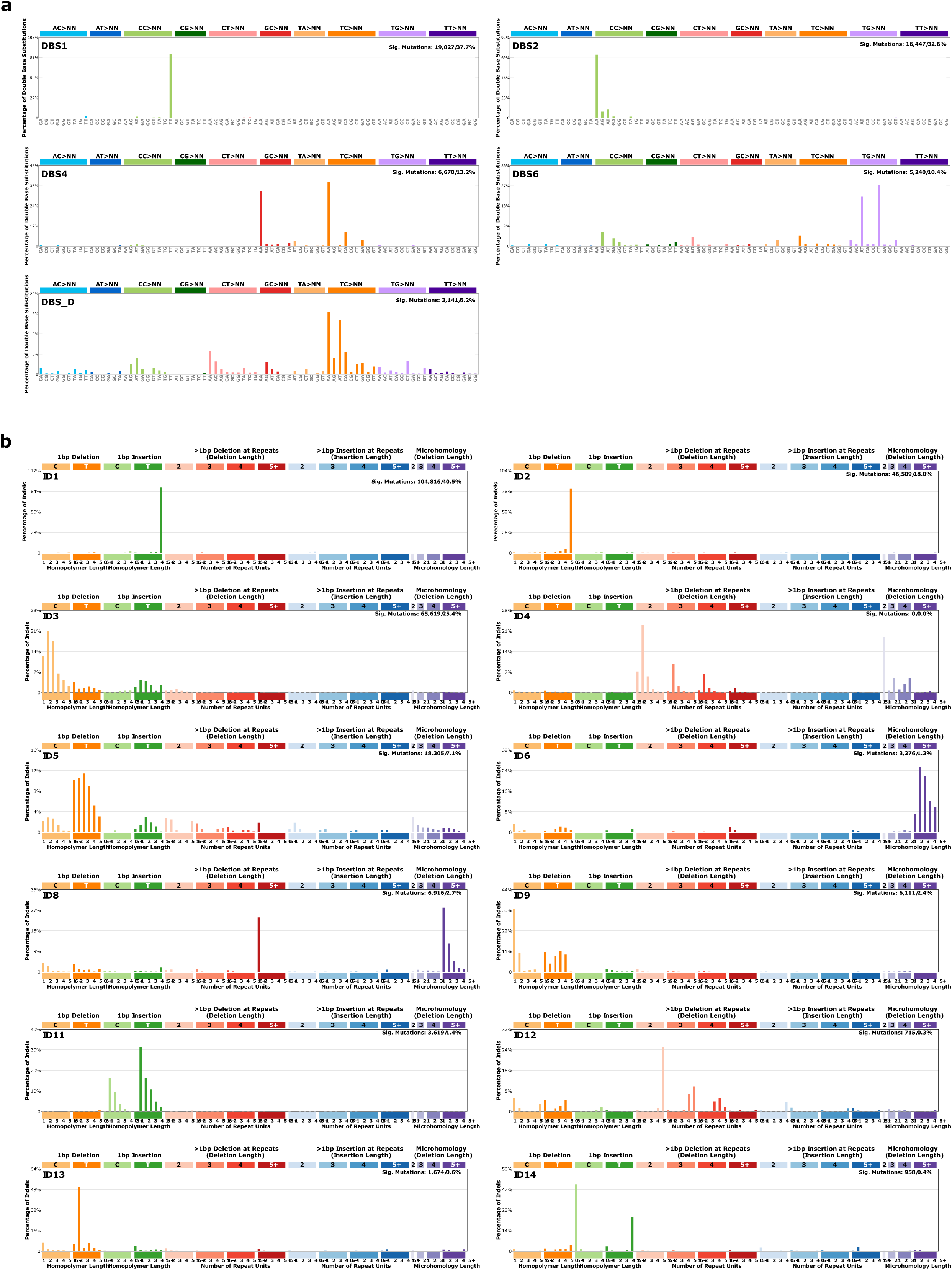
DBS and ID signature decomposition. Decomposed DBS (**a**) and ID (**b**) signatures, including reference COSMIC signatures and *de novo* signatures not decomposed into COSMIC reference signatures.

**Extended data Figure 4.**
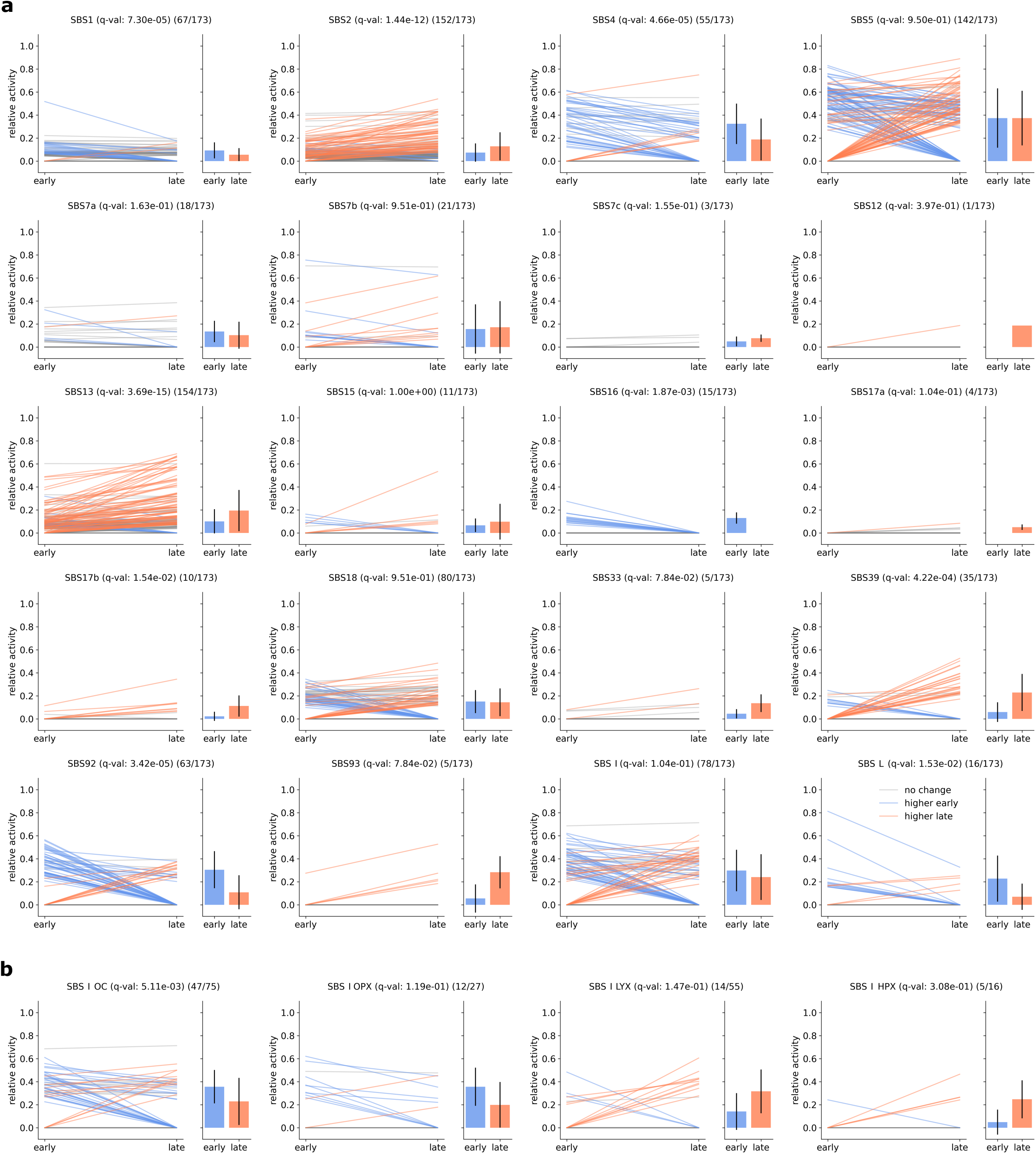
Evolutionary analysis of mutational signatures and driver mutations in HNC. **a**, Comparison of mutational signatures between early and late clonal mutations in HNC (*n*=173). **b,** Relative activities of SBS_I in early and late clonal mutations across anatomical subsites. Lines show the change in relative activity between the early and late clonal mutations within a positive sample. Colored lines represent an activity change of more than 6% (blue indicates higher in the clonal early mutations; orange indicates higher in the clonal late mutations). Bar plots show the distribution of activities in samples where the signature was present in the early and/or late clonal mutations; the number of positive samples is represented in the title of each plot. Black bars indicate one standard deviation away from the mean. Significance was assessed using a two-sided Wilcoxon signed-rank test, and p-values were corrected using the Benjamini-Hochberg Procedure (q-value). OC, oral cavity; OPC, oropharynx; LYX, larynx; HPX, hypopharynx.

**Extended Data Figure 5.**
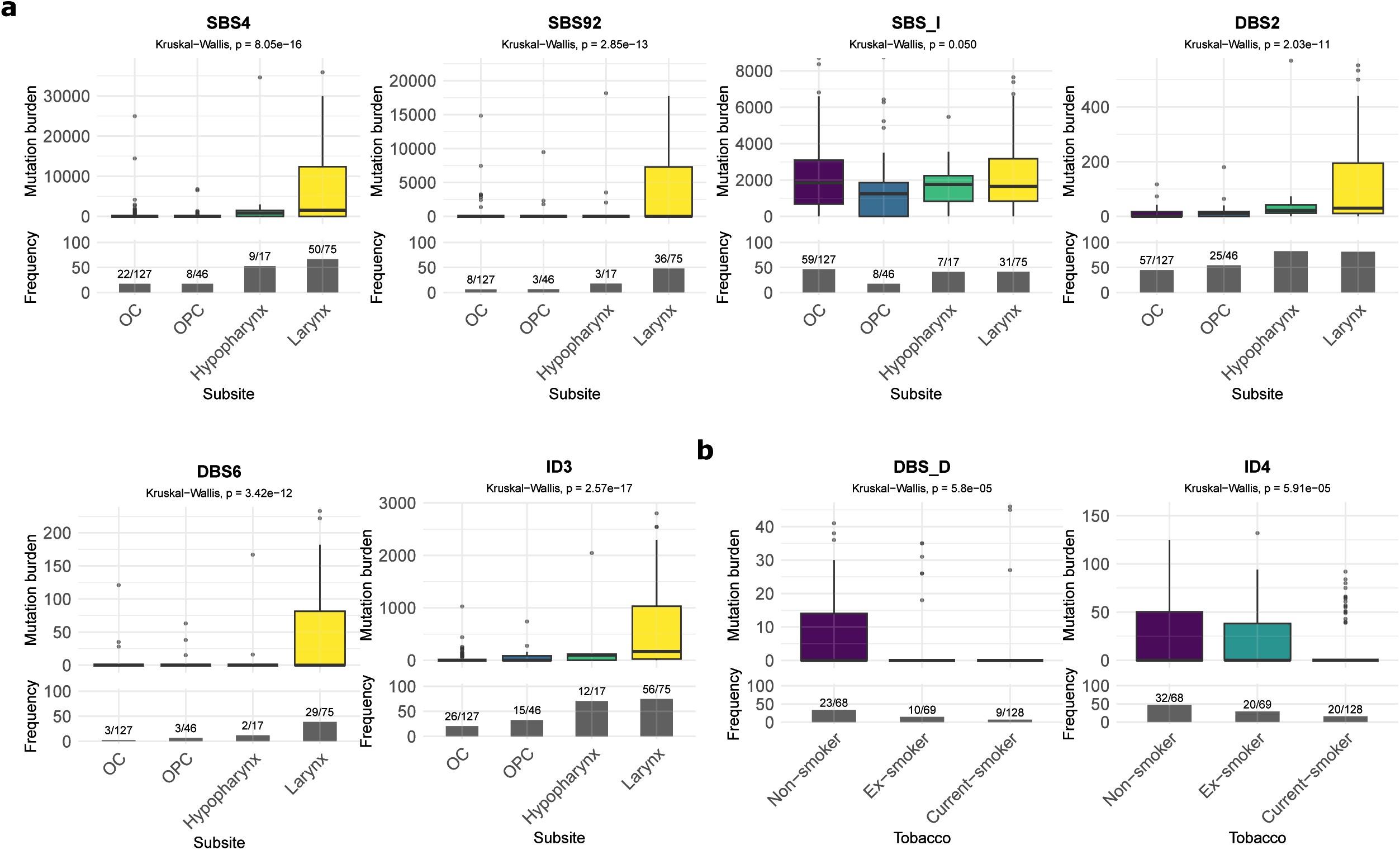
Association of tobacco-related mutational signatures with anatomical subsites and tobacco status. **a**, Mutational burdens for tobacco-related mutational signatures by anatomical subsite (n = 265 biologically independent samples). **b,** Mutational burdens for mutational signatures showing significant negative associations with tobacco consumption. The Kruskal–Wallis test (two sided) was used to test for global differences. Box-and-whisker plots are in the style of Tukey. The line within the box is plotted at the median, while upper and lower ends indicate 25^th^ and 75^th^ percentiles. Whiskers show 1.5 × interquartile range (IQR), and values outside it are shown as individual data points. Y axes were cut at 1.25 × upper whisker for clarity. Bar plots indicate the frequencies of dichotomized signaturesOC, oral cavity; OPC, oropharynx.

**Extended Data Figure 6.**
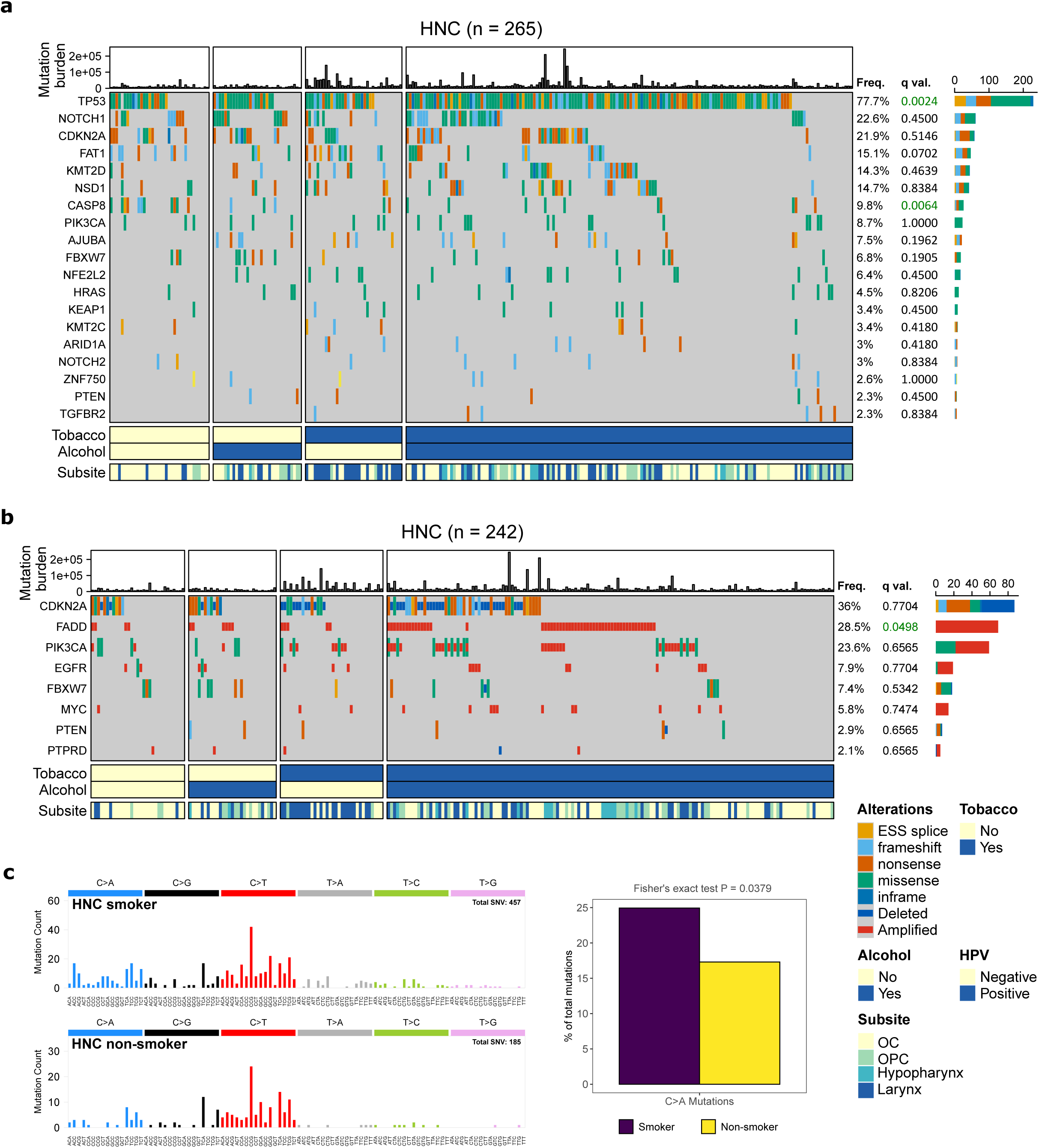
Driver alterations and driver mutation spectra in HNC. **a**, Driver mutations in HNC samples (*n*=265) sorted by tobacco and alcohol status. Genes mutated in more than 2% of the cases are shown. **b,** Driver mutations and copy number events in HNC samples with available copy number data (*n*=242). Only driver genes with both copy number gains and losses are included. Top, tumor mutational burden (TMB) per sample. Middle, presence of mutations per sample. Bottom, epidemiological characteristics. Frequency of mutations in the HNC dataset and q values from two-sided Fisher’s exact text are displayed. **c,** SBS96-mutation spectrum of driver mutations in smokers and non-smoker HNC cases and percentage of driver mutations occurring in C>A contexts.

**Extended Data Figure 7.**
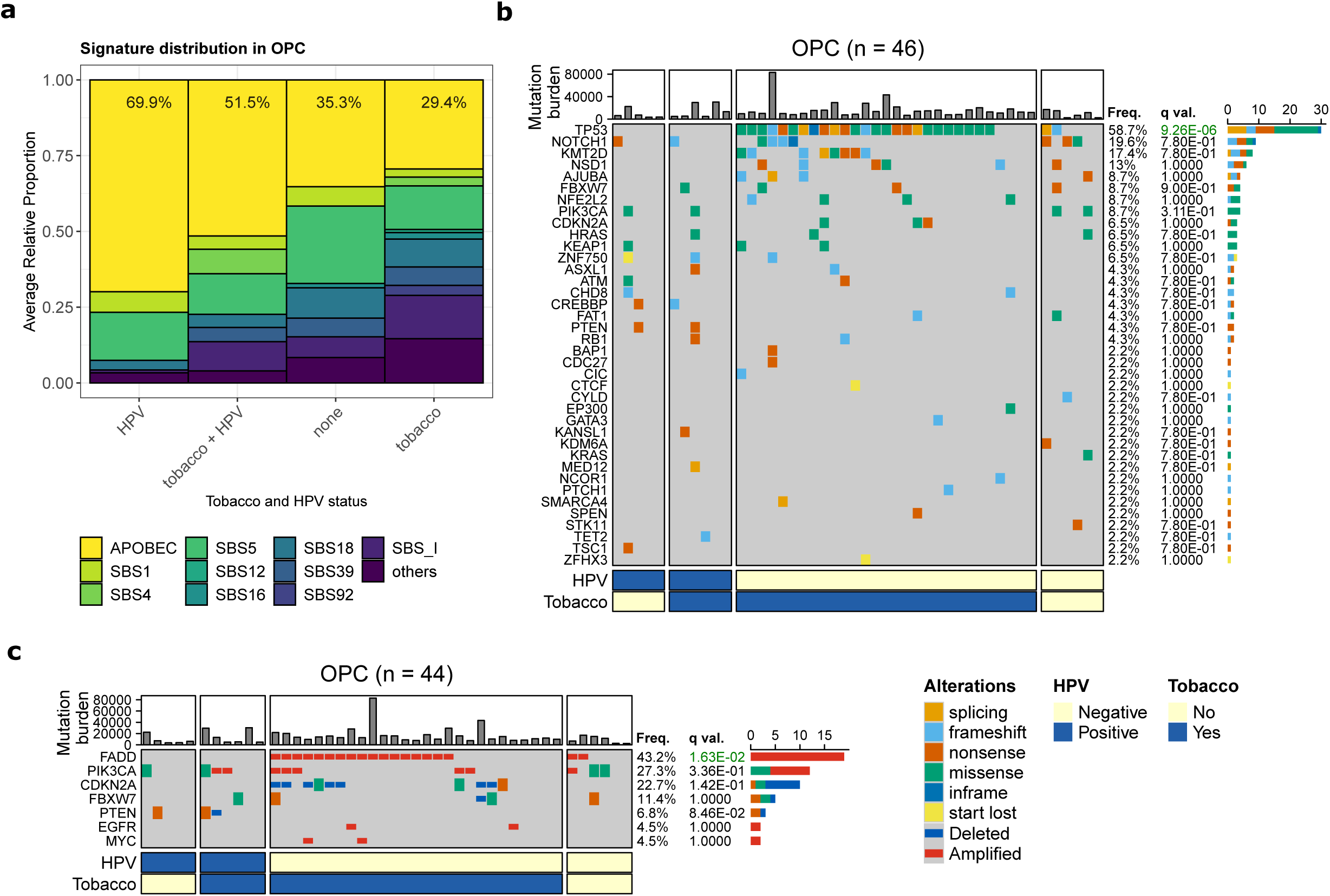
Mutational signature and driver spectra of oropharynx HNC cases by HPV status. **a**, Average relative attributions of SBS signatures by human papillomavirus (HPV) positivity and tobacco status in oropharynx (OPC) cancers. **b,** Driver mutations in OPC HNC samples (*n*=46) sorted by HPV and tobacco status. Genes mutated in more than 2% of the samples are shown. **c,** Driver mutations and copy number events in OPC HNC samples with available copy number data (*n*=44). Only driver genes with copy number gains and losses are included. Top, tumor mutational burden (TMB) per sample. Middle, presence of mutations per sample. Bottom, HPV status and tobacco smoking. Frequency of mutations in the HNC dataset and q values from two-sided Fisher’s exact text are displayed.

**Extended Data Figure 8.**
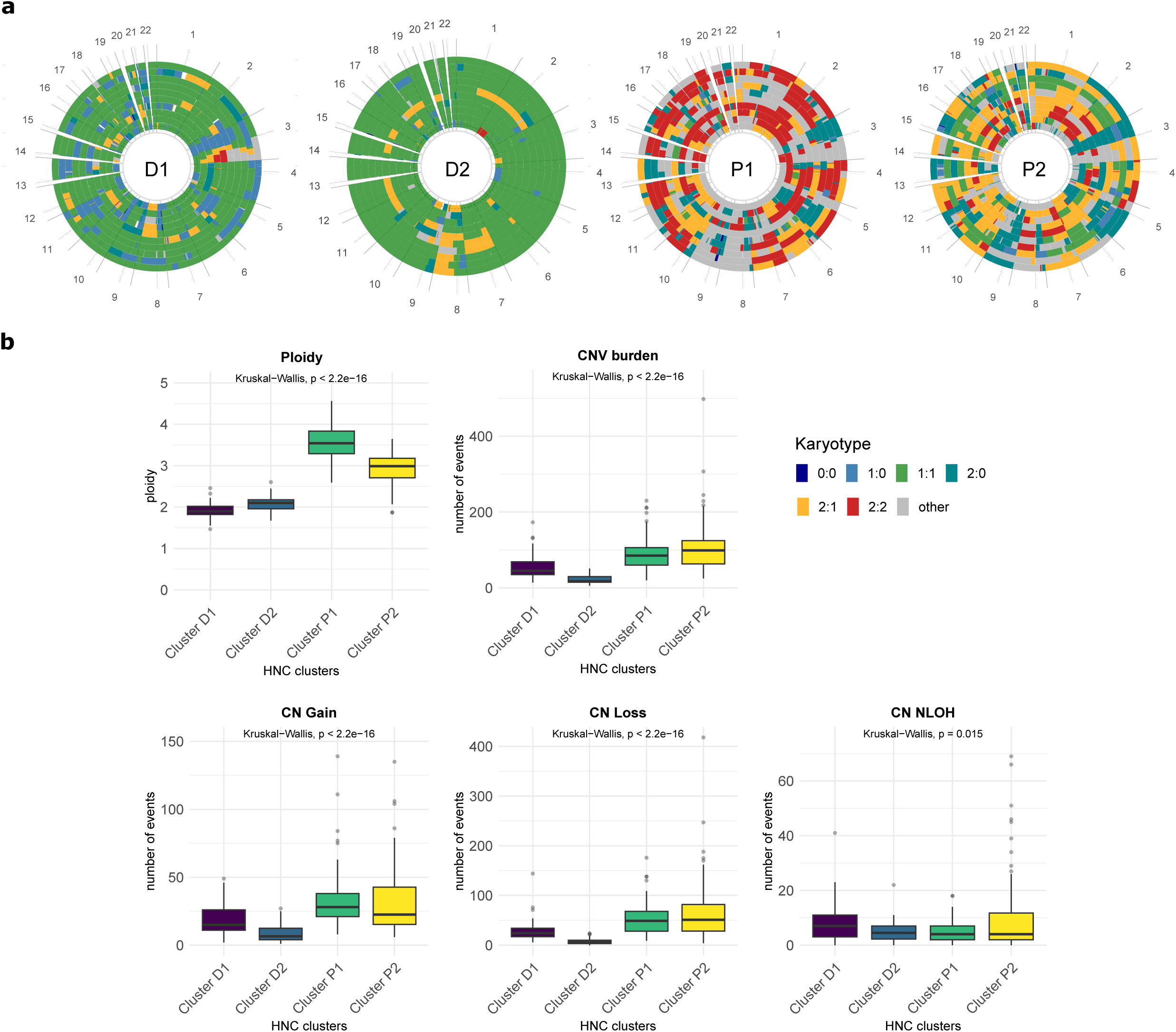
Copy number profile of head and neck cancer clusters. **a**, Genome-wide segments showing major and minor allele counts in 10 randomly picked samples per copy number (CN) cluster. **b,** Ploidy, CN burden, and burden of CN gains, losses, and CN neutral LOH (NLOH) across clusters (n = 242 biologically independent samples). The Kruskal–Wallis test (two sided) was used to test for global differences. Box-and-whisker plots are in the style of Tukey. The line within the box is plotted at the median, while upper and lower ends indicate 25^th^ and 75^th^ percentiles. Whiskers show 1.5 × interquartile range (IQR), and values outside it are shown as individual data points.

**Extended Data Figure 9.**
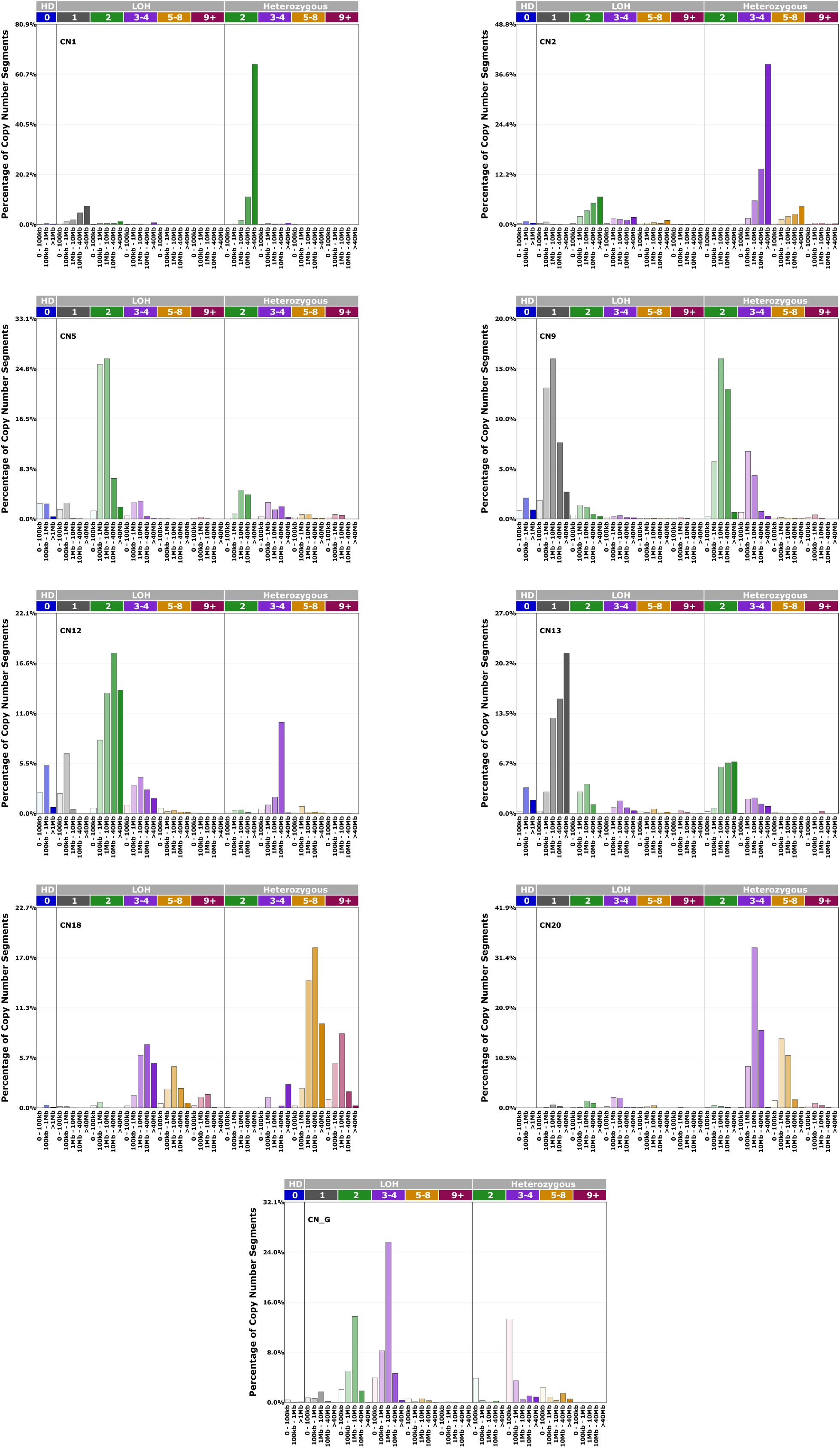
Copy number signature decomposition. Decomposed copy number signatures including reference COSMIC signatures and *de novo* signatures not decomposed into COSMIC reference signatures.

**Extended Data Figure 10.**
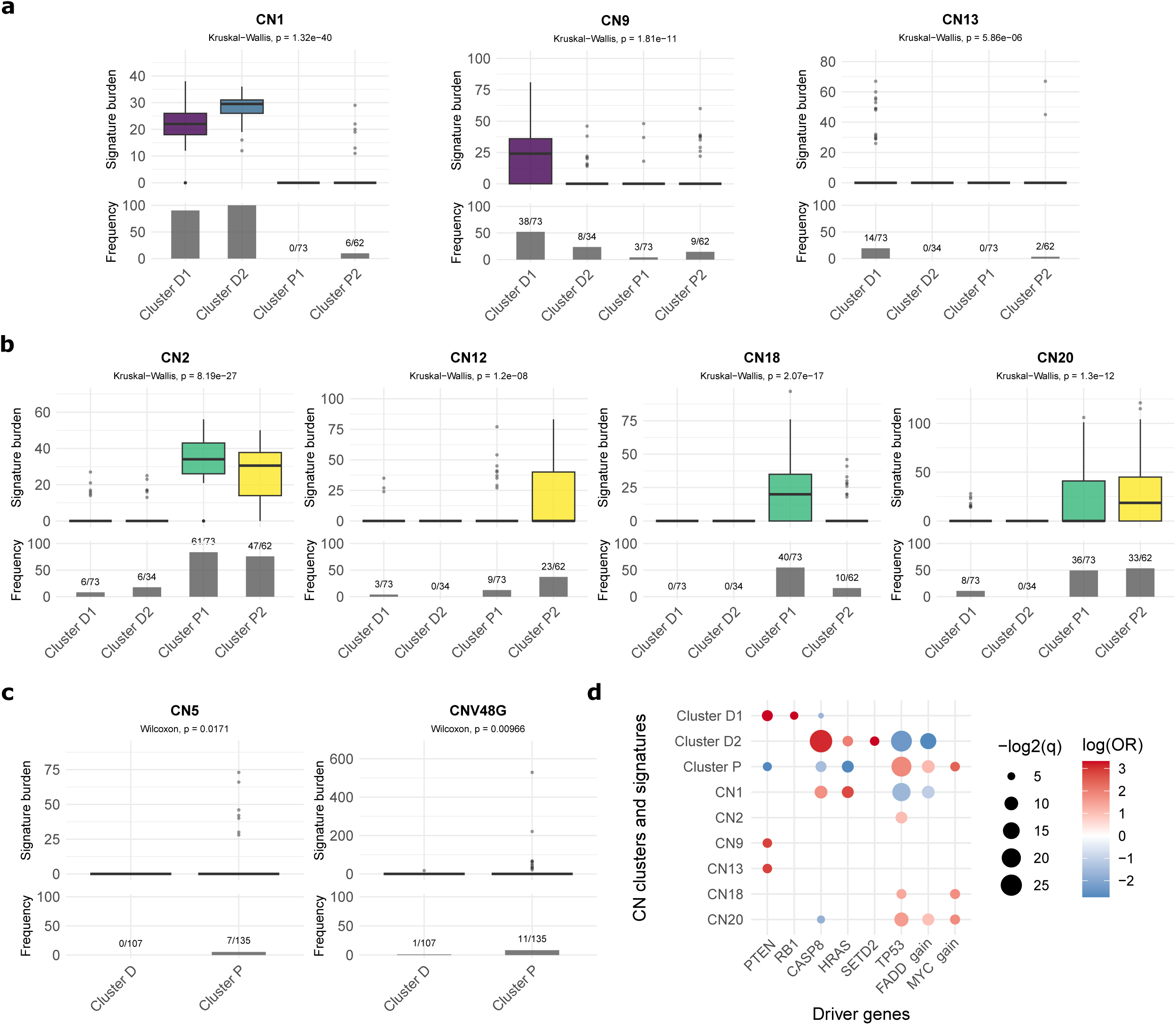
Copy number signature enrichment by HNC clusters and driver profile. **a-b**, Signature burdens for copy number signatures by copy number cluster showing associations with cluster D (**a**) and cluster P (**b**). **c**, Signature burdens for CN5 and CN_G signatures in copy number clusters D and P. The Kruskal–Wallis test (two sided) was used to test for global differences. Box-and-whisker plots are in the style of Tukey. The line within the box is plotted at the median, while upper and lower ends indicate 25^th^ and 75^th^ percentiles. Whiskers show 1.5 × interquartile range (IQR), and values outside it are shown as individual data points. Y axis were cut at 1.25 × upper whisker for clarity. Bar plots indicate the frequencies of dichotomized signatures. **d,** Associations between copy number clusters or signatures and driver alterations. Effect size (log2(OR), color), and significance level (–log2(q), size) from two-sided Fisher’s exact tests, corrected using the Benjamini-Hochberg Procedure, are displayed. Only significant associations are shown (q < 0.05).

## SUPPLEMENTARY TABLE AND FIGURE LEGENDS

**Supplementary Table 1. Clinical and epidemiological characteristics of the HNC dataset.**

**Supplementary Table 2. Mutation burdens per sample.**

**Supplementary Table 3. Associations of HNC risk factors with and mutational burden.**

**Supplementary Table 4. SBS *de novo* signatures extracted from 265 HNC cases.**

**Supplementary Table 5. DBS *de novo* signatures extracted from 265 HNC cases.**

**Supplementary Table 6. ID de *novo* signatures extracted from 265 HNC cases.**

**Supplementary Table 7. Decomposition of *de novo* mutational signatures to COSMIC reference signatures.**

**Supplementary Table 8. Activities of *de novo* mutational signatures in HNC cases.**

**Supplementary Table 9. Activities of decomposed COSMIC signatures in HNC cases.**

**Supplementary Table 10. Associations of HNC risk factors with COSMIC mutational signatures. Supplementary Table 11. Likely driver mutations identified in HNC.**

**Supplementary Table 12. Likely driver copy number alterations identified in HNC.**

**Supplementary Table 13. Associations of HNC risk factors with copy number clusters.**

**Supplementary Table 14. Details of individual case collections.**

**Supplementary Table 15. Details of data harmonization for HNC risk factors.**

**Supplementary Table 16. Details of HPV16 assessment in oropharynx cases.**

**Supplementary Table 17. CNV48 de novo signatures extracted from 242 HNC cases.**

**Supplementary Figure 1. Correlations amongst mutational signatures.** Pearson correlation coefficients for each significant comparison are indicated.

**Supplementary Figure 2. Association of tobacco-related signatures and HNC incidence.** Association between tobacco-related signatures with age-standardized rate (ASR) incidence. Number of mutations attributed to SBS4, SBS92, SBS_I, DBS2, DBS6, and ID3 mutational signatures against ASR of HNC per country, sex, and subsite. The p-values shown are for ASR variable in regressions across all cases, adjusted for age. The frequency of the signatures and number of cases per group are indicated.

**Supplementary Figure 3. Hierarchical clustering of copy number data.** Unsupervised hierarchical clustering analysis of copy number counts in the HNC cohort (*n*=242) using Euclidean distance and Ward’s agglomerative procedure. Two main clusters (diploid (D) and polyploid (P)) were obtained, which further subdivided into four groups. Right panel shows the copy number frequency in the HNC cohort.

**Supplementary Figure 4. Single base substitution signatures extracted by SigProfilerExtractor.** All single base substitution (SBS) de novo signatures extracted in SBS-1536 (15 signatures) and SBS-288 (14 signatures) format, shown side by side for comparison. Equivalent signatures where not extracted in SBS-288 format for SBS1536J. For clarity, the signatures context is retained in the signature names in this figure.

**Supplementary Figure 5.** Single base substitution mutational signatures extracted by mSigHdp. Fifteen single bases substitution (SBS) de novo signatures extracted by mSigHdp.

**Supplementary Figure 6.** Small insertion and deletion mutational signatures extracted by mSigHdp. Eight small insertion and deletion (ID) de novo signatures extracted by mSigHdp.

**Supplementary Figure 7. Mutational spectra supporting non-decomposed mutational signatures.** Individual mutational spectra are shown for cases which support the existence of non-decomposed signatures SBS_I (SBS1536I) (**a**), SBS_L (SBS1536_L) (**b**) and DBS_D (DBS78D) (**c**).

**Supplementary Figure 8. Principal component analysis of HNC SBS96 mutation counts and signature attributions.** Principal component analysis (PCA) performed on 256 cases of HNC on relative SBS96 mutation counts colored by **a,** anatomic site, **b,** tobacco status, and **c,** relative proportion of each mutation class (C>A, C>G, C>T, T>A, T>C, T>G). Circled on the anatomic site/ C>T plot is a subset of oral cavity HNC which have UV exposure. PCA performed on 256 cases of HNC on relative signature attributions colored by **d,** anatomic site, **e,** tobacco status and **f,** relative attributions of tobacco associated signatures SBS4, SBS92 and SBS_I.

**Supplementary Figure 9. UV exposure in HNC.** Support for the presence of UV in HNC of the oral cavity showing **a**, representative HNC oral cavity mutational specter which is consistent with representative melanoma mutational spectra from the PCAWG cohort and **b**, correlation between mutational signatures known to be associated with UV exposure in HNC. Correlation coefficients for each significant comparison are indicated.

**Supplementary Figure 10. Attribution of HNC mutational signatures in external datasets.** Attribution of HNC mutational signatures SBS_I (**a**) and SBS_L (**b**) in external datasets. The Kruskal– Wallis test (two sided) was used to test for global differences. Box-and-whisker plots are in the style of Tukey. The line within the box is plotted at the median, while upper and lower ends indicate 25th and 75th percentiles. Whiskers show 1.5 × interquartile range (IQR), and values outside it are shown as individual data points. Overall mutational signature landscape in in the external datasets was similar (**c**) with the presence of additional individual mutational spectra (**d**) supporting the existence of SBS_I.

**Supplementary Figure 11. Correlations amongst copy number signatures.** Pearson correlation coefficients for each significant comparison are indicated.

**Supplementary Figure 12. Predicted ancestry in HNC. a,** Scatter plots of principal components PC1 and PC2 based on genotype data showing the genetic structure of the HNC cohort across different countries of origin. **b,** Ancestry admixture in the HNC cohort. **c,** Probability of African ancestry by country.

**Supplementary Figure 13. Clustered mutations in HNC.** a-b, Distribution of clustered mutations in HNC by tobacco status (a) and anatomical subsite (b) ordered by median tumor mutational burden (TMB). Each dot represents a single tumor. The clustered mutation ratio is calculated as the fraction of clustered mutations compared to the total number of mutations in a given sample. Each clustered event is subclassified and summarized as the proportion of mutations per country associated with a double-base substitution event, an omikli event, or as a kataegis event.

**Supplementary Figure 14. Evolutionary analysis of driver mutations in HNC.** Relative frequency of driver mutations across early clonal and late clonal/subclonal stages, for the most common driver genes in HNC (*n*=173).

**Supplementary Figure 15. Human papillomavirus integration in HNC tumors. a,** Frequency of HPV16 integrations in genic and non-genic regions. **b,** Integration sites detected in chromosomal, cytoband, and genic regions. Rows represent samples positive for viral integration. The number of integrations per site and sample is depicted. Only four samples presented integrations in genic regions. **c-d,** Circos plots representing viral integration sites, structural variations (SV) and copy number (CN) alterations in tumor genomes presenting HPV16 integration. HPV integrations (in yellow) are depicted in the outermost ring, CN in the inner ring, and SV events in the center. Specific SV and CN alterations (CNA) surrounding the sites of integration (dotted lines) are shown for three samples (**d**).

**Supplementary Figure 16. Associations between germline *ADH1B* and *ADH7* variant genotype and alcohol related mutational signatures.** *ADH1B* rs1229984 and *ADH7* rs1573496 germline variant genotypes for signatures SBS16 (a), DBS4 (b) and ID11 (c) (*n*=265 biologically independent samples). Mutated samples correspond to those with at least one alternative allele. The Kruskal–Wallis test (two sided) was used to test for global differences. Box-and-whisker plots are in the style of Tukey. The line within the box is plotted at the median, while upper and lower ends indicate 25th and 75th percentiles. Whiskers show 1.5 × interquartile range (IQR), and values outside it are shown as individual data points. Y axes were cut at 1.25 × upper whisker for clarity. Bar plots indicate the frequencies of dichotomized signatures.

