## Supplementary Note for "The Complexity of Tobacco Smoke-Induced Mutagenesis in Head and Neck Cancer"

#### SUPPLEMENTARY INFORMATION

**This file contains the following:**

Supplementary Note

Supplementary References

Supplementary Figures

#### SUPPLEMENTARY NOTE

##### A) Supplementary Methods

**Extraction of *de novo* mutational signatures with SigProfilerExtractor**

For single base substitutions (SBS) extractions were performed with both SBS-288 and SBS-1536 contexts. SBS-288 extends the SBS-96 contexts by classifying mutations into transcribed, untranscribed, or intergenic non-transcribed regions whereas SBS-1536 considers the two flanking bases on either side of the mutated base to form a pentanucleotide context. These extractions extracted 14 and 15 signatures respectively (**Supplementary Figure 4**). Fourteen signatures were found in both extractions with high cosine similarity when collapsed to SBS96 contexts (**Supplementary Figure 4; Supplementary Note Table 1**).

The additional signature found in the SBS-1536 extraction was SBS1536J/SBS_J which was highly similar to SBS1536C / SBS_C (cosine similarity=0.98). These two signatures are both found in samples from the oral cavity which show evidence of ultraviolet light exposure and the additional signature is likely the result of overfitting of multiple samples with high mutation burdens. However, since both de novo signatures decompose readily into the Catalogue of Somatic Mutations in Cancer (COSMIC) reference signatures, this did not cause any issues with the downstream analysis.

**Extraction of *de novo* mutational signatures with mSigHdp**

In order to validate the mutational signatures obtained using SigProfilerExtractor, extractions were also performed with a second algorithm, mSigHdp[1] which is based on a hierarchical Dirichlet process as opposed to the nonnegative matrix factorization (NMF) approach utilised by SigProfilerExtractor. Unlike SigProfilerExtractor, the performance of mSigHdp has not been evaluated for extended contexts, therefore SBS extractions were only performed using SBS96 contexts. In total mSigHdp extracted 15 signatures, of which 13 were very close matches to the SBS1536 results, whereas hdp6 and hdp15 were unique to mSigHdp (**Supplementary Figure 5**). mSigHdp extracted eight indel signatures in comparison to the seven extracted from SigProfilerExtractor (**Supplementary Figure 6**), overall, the results were similar with mSigHdp extracting an additional signature containing ID1, ID2, and ID5. All Indel signatures in both extractions decomposed into COSMIC reference signatures.

**Decomposition to reference signatures**

The extracted de novo signatures were decomposed into the COSMIC reference signatures using SigProfilerAssignment (https://github.com/AlexandrovLab/SigProfilerAssignment; **Supplementary Table 7**).

On the issue of differences between SigProfilerExtractor and mSigHdp, hdp6 decomposes into SBS1, SBS5, SBS7a, and SBS18. All these signatures were also within the signature panel resulting from the decomposition of SigProfilerExtractor signatures, and no association was found with the de novo version of the signature. The other mSigHdp unique signature (hdp15), was not decomposed. However, since the signature could not be replicated in the SigProfilerExtractor results this signature was not included in the final signature panel.

**Justification for non-decomposed signatures**

Using default parameters, SigProfilerAssignment will reject the decomposition of a signature if the cosine similarity of the reconstructed signature is less than 0.8. Due to the large number of COSMIC reference mutational signatures, it is possible that the decompositions will include signatures which are implausible given the cohort type. As previously described[2], there are circumstances where it is justified to reject the decomposition result where the result is not plausible and where there is additional evidence to suggest that a mutational signature does represent a distinct mutational process.

SBS_I was originally decomposed to SBS1, SBS5, SBS8 and SBS85 with a cosine similarity 0.946. While SBS1, SBS5 and SBS8 are all reasonable, SBS85 is a signature associated with activation-induced cytidine deaminase (AID) which has only been found previously in lymphoid cancers, making this solution unlikely. Although there is no sample which has SBS_I as the dominant mutational process, it is possible to see evidence of the signature in individual mutational spectra (**Supplementary Figure 7a**). This, along with the observed association with tobacco smoking and oral cavity is more suggestive of a novel mutational signature. It is however likely that a more refined signature could be obtained from a cohort of either larger size or with individuals with higher mutational burden from this signature.

For SBS_L, the original decomposition included SBS1, SBS5, SBS30 and SBS44 with a cosine similarity of 0.889. The combination of these signatures is unlikely, as SBS30 is a consequence of *NTLH1* mutations and SBS44 which is a signature associated with microsatellite instability (MSI). MSI cases would typically show large numbers of indels, which is not the case for samples where SBS_L is attributed, and neither were *NTLH1* mutations present. Like SBS_I there are no samples which have SBS_L as the dominant mutational process. The samples with the highest relative attribution to this SBS_L (>50% of the total mutation burden) also have a high level of attribution to APOBEC-associated signatures 2/13 (**Supplementary Figure 7b**). As SBS2 overlaps in the T[C>T]N contexts it is possible that these two signatures could not be completely separated during the extraction process. Similar to SBS_I, larger cohorts or individuals with higher mutation burdens from this signature may yield an improved version of this signature.

DBS_D is the only other signature which remains non-decomposed, however, in this case the decomposition is rejected automatically by SigProfilerAssignment as the cosine similarity of the reconstructed signature is only 0.727. In support of this there is a sample where DBS_D is the dominant signature in the mutational spectra (**Supplementary Figure 7c**), in addition to the observed enrichment in non-smokers.

**Analysis of clustered mutations**

Clustered mutations were classified and analyzed using SigProfilerClusters (v1.1.2), which is designed to filter out clustered mutations from complete somatic mutational catalogs[3]. Specifically, SigProfilerSimulator (v1.1.5) was used to derive an inter-mutational distance (IMD) cutoff that is unlikely to occur by chance given the mutational patterns and tumor mutational burden of each sample[4]. Each sample was simulated while maintaining the +/-2bp sequence context and the transcriptional strand bias ratios across all mutations. All samples were simulated 100 times with the IMD cutoff being calculated in which 90% of all mutations below the distance cutoff do not appear by chance (q-value<0.01). P-values were calculated using z-tests by comparing the distribution of simulated mutations to the number of real mutations occurring within the same IMD cutoff and were corrected for multiple hypothesis testing using Benjamini-Hochberg FDR procedure. A maximum threshold of 10 kb was used for all IMD cutoffs. Heterogeneity in mutation rates across the genome was also considered by correcting for mutation-rich regions present in 1Mb-sized windows. Further, the effects of clonality and copy number were addressed using a threshold for the difference in variant allele frequencies (no more than 0.1) to ensure that a given subset of mutations is likely to have occurred as a single event. The subsequent clustered mutations were subclassified into specific categories of mutational events consisting of i) doublet-base substitutions, reflecting two adjacent mutations with consistent variant allele frequencies; ii) extended multi-base substitutions, previously termed omikli events[5], reflecting two or three mutational events with at least a single IMD greater than 1bp and less than the sample-dependent IMD threshold with consistent variant allele frequencies; iv) large mutational events, previously termed kataegis[6], reflecting four or more mutational events with at least a single IMD greater than 1bp and less than the sample-dependent IMD threshold with consistent variant allele frequencies[7].

**Germline variant calling**

The WGS-based germline genotype data of 265 HNC cases were processed as described elsewhere[2]. Briefly, germline variants from paired blood samples of HNC cancer patients were jointly called by the gVCF gvcfgenotyper tool (version: master_2019.02.26). In the quality control steps using PLINK (v1.9b, www.cog-genomics.org/plink/1.9/), we kept 11,605,626 biallelic variants with genotype missing rate <10% that did not fail Hardy-Weinberg equilibrium test (p<1e-08). Variant calls were then derived into genotypes for each individual and annotated using SnpEff (version 5.0) from the dbSNP database (version 150). Genotypes for variants related to HNC risk were extracted for each case.

**Genomic ancestry and admixture analyses**

We used ADMIXTURE tool (v1.3.0)[8] to infer the genetic ancestry of individuals within HNC cases. The admixture and principal component analyses were restricted to Hapmap SNPs. We additionally excluded germline variants with minor allele frequency <1% within regions of long-range, high linkage disequilibrium in the human genome (hg38), remaining 1,182,596 variants. After pruning for linkage disequilibrium (r2<20% within 50kb window), 159,464 independent variants remained in HNHNC genotype data. The 1000 genome reference population genotype data (phase 3) for Europeans (*n*=489), Africans (*n*=661), East Asians (*n*=504), and Latin Americans (N=347) (https://www.internationalgenome.org/data/) were filtered and merged with HNC genotype data based on the pruned set of variants present in both datasets. Admixture analysis was performed on the merged genotype data with K=4, which would correspond to the four ancestral continental population groups that would reflect the participants of our study. To complement the Admixture results, principal component analysis was applied to the same samples and HNC cases were visualized in two dimensions in comparison with each reference population included in the 1000 genome dataset.

**Droplet digital PCR MSI assay**

The presence of MSI in HNC tumour samples was assessed using the QX200 Droplet Digital PCR System (Bio-Rad, Hercules, CA, USA) for the detection of five microsatellite (MS) markers (BAT25, BAT26, NR21, NR24, Mono27) commercially pooled in three primer–probe mix assays, as previously described[9]. Briefly, samples were tested in duplicate, and each reaction comprised 1× ddPCR Multiplex Supermix for probes (Bio-Rad), 1X primer–probe mix, and 10 ng of extracted tumor DNA, in a total volume of 22 µl. MSI-positive, negative, and no-template (nuclease-free water) controls were included in each experiment. Droplet generation and plate preparation for thermal cycling amplification were performed using the QX200 AutoDG Droplet Digital PCR System (Bio-Rad). The following PCR protocol was applied on a C1000 Touch Thermal Cycler (Bio-Rad): 37 °C for 30 min, 95 °C for 10 min, followed by 40 cycles of denaturation at 94 °C for 30 s, annealing at 55 °C for one minute, with a final extension at 98 °C for 10 min. Following PCR amplification, fluorescence signals were quantified using the QX200 Droplet Reader (Bio-Rad), and data were analysed with QuantaSoft Analysis Pro v.1.0.596.0525 (Bio-Rad) software. Positive and negative controls served as guides to call markers and delineate clusters. For each assay, the cluster at the bottom left of the x–y plot was designated as the negative population. Clusters located vertically and horizontally from the negative cluster were identified as the mutant population, while clusters located diagonally from the negative cluster represented the wild-type population. Tumors were characterized for the MSI phenotype by analyzing the results for all five markers using the following criteria: MSI if two or more mutant MS markers were observed, and MSS when none or only one of the MS markers was altered.

**B) Supplementary Results**

**Principal component analysis of mutatin SBS96 counts and SBS signature attributions**

PCA analysis was performed both on both the relative SBS96 mutation counts and relative SBS signature attributions (**Supplementary Figure 8**). For the SBS96 mutation counts there was a clear separation of a subset of cases which were predominantly smokers from the larynx (**Supplementary Figure 8a-b**). Coloring the PCA plots by the relative proportions of the 6 major mutation classes (C>A, C>G, C>T, T>A, T>C and T>G, **Supplementary Figure 8c**) shows that this subset of larynx/smokers is characterized by more C>A mutations, which would be consistent with mutations caused by the tobacco-associated mutational signature SBS4. The PCA also shows separation of a subset of cases with higher relative proportion of C>G and C>T mutations. A small portion of this subset consists of oral cavity HNC with UV exposure, as shown by the higher relative proportion of C>T mutations in this group while the remainder of the subset corresponds to cases that have high levels of APOBEC activity (**Supplementary Figure 8a,c**). As APOBEC is not found exclusively in non-smokers this likely explains why there is not a complete separation of smokers from non-smokers in the PCA analysis.

PCA analysis of the relative signature attributions also shows separation of a subset of larynx/smoker cases (**Supplementary Figure 8d-e**). Coloring the plots by relative attribution of the tobacco-associated signatures confirms that this subset is defined by high contributions from SBS4 and SBS92, but not SBS_I (**Supplementary Figure 8f**). Analysis of additional PCA analysis components for both SBS96 mutation counts and signature attribution did not reveal any additional insights other than a greater separation of the UV-exposed subset of cases with PC3/4. These results taken together provide additional support for the tissue specificity observed in tobacco-associated mutational signatures, as these differences are clear even when using raw mutation counts prior to extraction of mutational signatures.

**Presence of UV-related signatures**

The finding of SBS7 in oral cavity HNC other than those in the lip was surprising, considering that these tissues are not external. In order to provide confidence in the presence of SBS7 in these cases, a number of additional tests were performed. Firstly, the cases were reviewed by expert pathologists to confirm the classification as HNC tumors of the inner lip (*n*=3), unspecified lip (*n*=1), tongue (*n*=2), and floor of the mouth (*n*=7) as opposed to the external lip. Secondly, the mutational spectra of positive cases were reviewed to confirm that the profile was consistent with UV exposure when compared to UV-exposed skin cancers from the PCAWG cohort (**Supplementary Figure 9a**). Thirdly, we checked for correlations between the SBS signatures SBS7a/b/c and other signatures of UV exposure, with a strong correlation found with both DBS1 and ID13 as expected (**Supplementary Figure 9b**)[10]. Taken together, these tests provide confidence that UV exposure is present in oral cavity HNC.

**Attribution of mutational signatures in external data sets**

In order to confirm whether the newly extracted signatures were found in an independent cohort, we attributed our panel of signatures using SigProfilerAssignment to both our samples and two cohorts of HNC samples from the PCAWG data set from the US (PCAWG-HNSC, *n*=39) and India (PCAWG-ORCA, *n*=13)[11]. SBS_I was present in 9/39 (23%) of the samples from the US but only 1/13 (7%) of the samples from the Indian cohort. Whilst this could be due to the small cohort size it is also worth noting that SBS_I was associated with tobacco exposure, and the Indian cohort has a higher proportion of non-smokers compared to both the US cohort and the cohort in this study (5/13 (38.5%) in PCAWG-ORCA vs 9/39 (23.1%) in PCAWG-HNSC and 68/265 (25.7%) in MUTOGRAPHS). All cases positive for SBS_I were oral cavity HNC, which is consistent with the enrichment of SBS_I in oral cavity HNC observed in this study (**Supplementary Figure 10a,c**). In addition, SBS_I can be observed in individual mutational spectra from the PCAWG cohorts (**Supplementary Figure 10d**). SBS_L was not found in any of the PCAWG cases, although this signature was much rarer in our cohort compared to SBS_I (**Supplementary Figure 10b-c**). Finally, in the PCAWG cohort, SBS4 and SBS92 were found in the highest proportions in the larynx, consistent with the observation that both signatures were enriched in this anatomic site (**Supplementary Figure 10c**).

**Smoking and drinking habits and link to mutation profile**

We evaluated the associations between additional smoking parameters with mutation and tobacco-related signature burdens, including tobacco quantity, defined as cigarettes per day, and smoking duration, defined as years smoking. Linear regressions were performed with signature attributions with confidence intervals not consistent with zero or mutation burdens as a dependent variable. Sex, age of diagnosis, subsite, region, and alcohol status were added as covariates. All tobacco-related signatures (SBS4, SBS92, SBS1536I, ID3, DBS2, DBS6), as well as total mutation burdens, were positively correlated with the quantity of cigarettes and smoking duration (**Supplementary Note Table 2**). This suggests that the amount of tobacco consumed, rather than just the presence of the risk factor, influenced the accumulation of tobacco-related mutations in HNC.

We also investigated whether the quantity of alcohol consumed (in grams per week) could act as a confounding variable in the association between alcohol-related signatures (SBS16, DBS4, ID6, and ID11) and tobacco plus alcohol status. Specifically, we ran a logistic regression with binary attributions as dependent variables and tobacco plus alcohol status as independent variable (Methods). We included alcohol quantity, sex, age of diagnosis, subsite, and region as covariates. The association between these signatures and the combined tobacco and alcohol exposure status remained significant, thereby confirming that the results are not confounded by higher alcohol consumption among tobacco smokers (**Supplementary Note Table 3**). Associations with alcohol quantity were not significant for any signature.

**Copy number signatures extracted in HNC**

To unveil distinct CN particularities within each CN cluster and etiology, we conducted CN signature extraction[12]. Our analysis yielded 7 *de novo* CN signatures, which collapsed into 8 COSMIC reference signatures, along with one non-decomposed signature, CN_G (**Extended Data Figure 9; Supplementary Table 17; Supplementary Note Tables 4-6**). Signatures of diploidy (CN1) and tetraploidy (CN2) were present in 44% and 50% of the samples, respectively, and exhibited negative correlation (**Figure 7a; Supplementary Figure 11**). We also identified signatures related to loss of heterozygosity on diploid backgrounds (CN9 in 24.0% of the samples and CN13 in 6.6%, respectively), and in the context of whole-genome duplication (CN12 in 14.5%). Two signatures of unknown etiology, CN18 and CN20, characterized by complex CN patterns with double and single whole-genome duplication, were detected in 20.7% and 14.5% of cases, respectively. Finally, chromothripsis signature CN5 and signature CN_G, capturing an uncharacterized genome instability profile, were observed in small fractions of samples (2.9% and 5.0%, respectively).

**Copy number signature of chromosomal instability in Brazil**

The previously unextracted CN signature CN_G was present exclusively in HNC cases from South American patients (8% [12/150] vs 0% [0/92] in Europe, p=0.0041). Specifically, CN_G was detected in samples from Brazil and Argentina (8.8% [11/125] and 4.4% [1/23], respectively). To further investigate this, we explored the genetic ancestry of patients included in our dataset (**Supplementary Methods**) and we observed a positive association between the likelihood of African ancestry and CNV48 burden, as well as total CN burden (**Supplementary Note Tables 7-8; Supplementary Figure 12**). This aligns with previous reports indicating higher genome instability in HNC cases with African ancestry and suggests that CN48G could recapitulate the enhanced chromosomal instability that is prevalent in this population[13,14]. However, it cannot be ruled out that a potential misalignment of genomes of African descent with reference genomes could be responsible for the observed associations. No association was found between genetic ancestry profiles and other signatures (**Supplementary Note Table 8**).

**Clustered mutations**

We investigated the number and type of clustered mutations in HNC tumors, including doublet base substitutions (DBS), omikli events[5], and kataegis events[6] (**Supplementary Methods**). The burden of clustered mutations out of the total mutations was significantly higher in HNC cases from smokers compared to non-smokers (Kruskal-Wallis test, p=4.72E$^{-4}$, **Supplementary Figure 13a**). Most clustered mutations in HNC from smokers corresponded to DBS, in line with the high burdens of tobacco-related DBS2 found in these samples (**Figure 3**). Conversely, clustered events among non-smokers corresponded predominantly to omikli, which have previously been attributed to APOBEC3 activity[5,7]. The distribution of clustered mutations also varied across anatomical subsites, with larynx cases presenting higher burdens (Kruskal-Wallis test, p=0.0075, **Supplementary Figure 13b**). Other epidemiological variables such as country of origin and alcohol status, did not present significant differences.

**Molecular timing of driver mutations**

The clonality of driver single-base substitutions annotated with MutationTimeR[15] in 174 tumor samples was retrieved. Driver mutations were classified as early clonal (*n*=285), late clonal (*n*=24), or subclonal (*n*=5). We observed an enrichment of *T53* mutations in clonal early stages compared to clonal late and subclonal stages, with 120 *TP53* driver mutations being annotated as early clonal (42.1% of clonal mutations) compared to three annotated as late clonal or subclonal (10.3%) (Fisher's test, p=0.0005; **Supplementary Figure 14**). Considering this, *TP53* drivers could play a role in genome instability during early evolutionary stages in positive samples[15], potentially contributing to the genomic profiles observed in cases exposed to tobacco and in the CN cluster P (**Figure 7d**).

**Mismatch repair deficiency assessment**

Attribution of COSMIC signature SBS15, related to defective DNA mismatch repair[16], was present in 53/265 samples (**Figure 2a**). In cases with microsatellite instability (MSI), SBS15 commonly contributes to large numbers of substitutions. Conversely, in the HNC dataset, SBS15 accounted for <10% of the mutation burden in all samples except for one (PD52800a). This sample was deemed microsatellite stable based on droplet digital PCR MSI assay (**Supplementary Methods**) and did not present deleterious germline or somatic mutations in mismatch repair genes (*MLH1*, *MSH2*, *MSH6*, *PMS1*, and *PMS2*). Overall, our analysis suggests that all samples in the HNC dataset presented a microsatellite-stable phenotype despite the presence of SBS15. This is consistent with the low frequencies of MSI commonly observed in HNC[17].

**HPV genome detection and integration**

Two NGS-based viral integration tools, VERSE and FastViFi, were used to detect the presence of HPV16 genomes and viral integration sites in oropharynx tumor samples[18,19]. Given VERSE is shown to be stringent at detecting integration sites[20], viral integration was determined by FastViFi. FastViFi relies on read-mapping to host, viral reference genome, and an ensemble of hidden Markov models (eHMMs) to identify viral integration sites[18,19]. Out of 10 oropharynx samples with detectable HPV16 genomes according to FastViFi, seven presented HPV16 integration (**Supplementary Table 16**). Integration sites were distributed stochastically in the genome, with no evident genomic clusters of viral integrations, and were more prevalent in non-genic regions, in line with previous studies (**Supplementary Figure 15a-b**)[20]. Recurring integration events in genic regions were not observed in our dataset, but we detected an integration within *STX17* and PLGRTK also described in previous studies[20].

Our results revealed structural variants surrounding the sites of integration. Specifically, HPV insertional breakpoints were found at regions of genomic amplification, deletion, or at the junction between chromosomal translocations, including a translocation event between two HPV integration sites (**Supplementary Figure 15c-d**). This association between insertion sites and structural variations aligns with the hypothesis that HPV16 infection may lead to genome instability through several proposed mechanisms, which may consequently facilitate viral integration[21].

**Germline variants related to HNC risk**

The presence of deleterious germline variants previously described in HNC was assessed to identify any associations with mutational signatures. Variants in alcohol dehydrogenase 1B (*ADH1B*) and alcohol dehydrogenase 7 (*ADH7*) have been associated with enhanced alcohol metabolism to acetaldehyde and low alcohol dependency[22,23]. In our dataset, the *ADH1B* variant rs1229984 was detected in 6.8% (18/265), and the *ADH7* variant rs1573496 in 11% (29/265), consistent with expected frequencies for the studied population[24]. Both variants demonstrated a significant association with a low burden of alcohol-related signatures, suggesting that patients carrying these protective variants present reduced alcohol-related DNA damage (**Supplementary Note Table 9; Supplementary Figure 16**). The *aldehyde dehydrogenase 2* (*ALDH2*) variant rs671, linked to poor alcohol metabolism and enhanced HNC risk[22,23], was also identified in one case.

Germline variants related to Fanconi anemia were also investigated. Fanconi anemia is a rare genetic disease caused by biallelic mutations in *FANC* genes, which lead to impaired DNA damage repair that confers an increased risk of developing HNC[25,26]. Variants with a putative deleterious impact were assessed using SnpEff and ClinVar annotations. One individual was heterozygote for a variant in *FASL*, but no heterozygous pathogenic mutation was identified in FANC genes, indicating that none of the cases in the HNC dataset presented the disease (**Supplementary Note Table 9**). Other pathogenic variants related to DNA repair genes were identified in *BRCA2* (*n*=2) and *ATM* (*n*=1), with no associations with the mutational signature profile.

#### SUPPLEMENTARY TABLE AND FIGURE LEGENDS

**Supplementary Note Table 1. Comparison of single base substitution signatures extracted by SigProfilerExtractor and mSigHdp.**

**Supplementary Note Table 2. Associations of cigarette quantity (cigarettes per day) and smoking duration (years) with mutation burdens and tobacco-related mutational signatures.**

**Supplementary Note Table 3. Associations of COSMIC mutational signatures with tobacco and alcohol status corrected by alcohol quantity.**

**Supplementary Note Table 4. Decomposition of de novo copy number signatures to COSMIC reference signatures.**

**Supplementary Note Table 5. Activities of de novo copy number signatures in HNC cases.**

**Supplementary Note Table 6. Activities of decomposed COSMIC copy number signatures in HNC cases.**

**Supplementary Note Table 7. Admixture analysis in HNC samples.**

**Supplementary Note Table 8. Associations of CN signatures with African ancestry.**

**Supplementary Note Table 9. Germline variants related to alcohol metabolism and DNA repair.**

**Supplementary Figure 1. Correlations amongst mutational signatures.** Correlation coefficients for each significant comparison are indicated.

**Supplementary Figure 2. Association of tobacco-related signates and HNC incidence.** Association between tobacco-related signatures and age-standardized rate (ASR) incidence. Number of mutations attributed to SBS4, SBS92, SBS_I, DBS2, DBS6, and ID3 mutational signatures against ASR of HNC per country, sex, and subsite. The p-values shown are for ASR variable in regressions across all cases, adjusted for sex. The frequency of the signatures and number of cases per group are indicated.

**Supplementary Figure 3. Hierarchical clustering of copy number data.** Unsupervised hierarchical clustering analysis of copy number counts in the HNC cohort (*n*=242) using Euclidean distance and

Ward’s agglomerative procedure. Two main clusters (diploid (D) and polyploid (P)) were obtained, which further subdivided into four groups. Right panel shows the copy number frequency in the HNC cohort.

**Supplementary Figure 4. Single base substitution signatures extracted by SigProfilerExtractor.** All single base substitution (SBS) de novo signatures extracted in SBS-1536 (15 signatures) and SBS-288 (14 signatures) format, shown side by side for comparison. Equivalent signatures where not extracted in SBS-288 format for SBS1536J. For clarity, the signatures context is retained in the signature names in this figure.

**Supplementary Figure 5. Single base substitution mutational signatures extracted by mSigHdp.** Fifteen single bases substitution (SBS) de novo signatures extracted by mSigHdp.

**Supplementary Figure 6. Small insertion and deletion mutational signatures extracted by mSigHdp.** Eight small insertion and deletion (ID) de novo signatures extracted by mSigHdp.

**Supplementary Figure 7. Mutational spectra supporting non-decomposed mutational signatures.** Individual mutational spectra are shown for cases which support the existence of non-decomposed signatures SBS_I (SBS1536I) (**a**), SBS_L (SBS1536_L) (**b**) and DBS_D (DBS78D) (**c**).

**Supplementary Figure 8. Principal component analysis of HNC SBS96 mutation counts and signature attributions.** Principal component analysis (PCA) performed on 256 cases of HNC on relative SBS96 mutation counts colored by **a,** anatomic site, **b,** tobacco status, and **c,** relative proportion of each mutation class (C>A, C>G, C>T, T>A, T>C, T>G). Circled on the anatomic site/ C>T plot is a subset of oral cavity HNC which have UV exposure. PCA performed on 256 cases of HNC on relative signature attributions colored by **d,** anatomic site, **e,** tobacco status and **f,** relative attributions of tobacco associated signatures SBS4, SBS92 and SBS_I.

**Supplementary Figure 9. UV exposure in HNC.** Support for the presence of UV in HNC of the oral cavity showing **a**, representative HNC oral cavity mutational spectre which is consistent with representative melanoma mutational spectra from the PCAWG cohort and **b**, correlation between mutational signatures known to be associated with UV exposure in HNC. Correlation coefficients for each significant comparison are indicated.

**Supplementary Figure 10. Attribution of HNC mutational signatures in external datasets.** Attribution of HNC mutational signatures SBS_I (**a**) and SBS_L (**b**) in external datasets. The Kruskal–Wallis test (two sided) was used to test for global differences. Box-and-whisker plots are in the style of Tukey. The line within the box is plotted at the median, while upper and lower ends indicate 25th and 75th percentiles. Whiskers show 1.5 × interquartile range (IQR), and values outside it are shown as individual data points. Overall mutational signature landscape in in the external datasets was similar (**c**) with the presence of additional individual mutational spectra (**d**) supporting the existence of SBS_I.

**Supplementary Figure 11. Correlations amongst copy number signatures.** Correlation coefficients for each significant comparison are indicated.

**Supplementary Figure 12. Predicted ancestry in HNC. a,** Scatter plots of principal components PC1 and PC2 based on genotype data showing the genetic structure of the HNC cohort across different countries of origin. **b,** Ancestry admixture in the HNC cohort. **c,** Probability of African ancestry by country.

**Supplementary Figure 13. Clustered mutations in HNC. a-b,** Distribution of clustered mutations in HNC by tobacco status (**a**) and anatomical subsite (**b**) ordered by median tumor mutational burden (TMB). Each dot represents a single tumor. The clustered mutation ratio is calculated as the fraction of clustered mutations compared to the total number of mutations in a given sample. Each clustered event is subclassified and summarized as the proportion of mutations per country associated with a double-base substitution event, an omikli event, or as a kataegis event.

**Supplementary Figure 14. Evolutionary analysis of driver mutations in HNC.** Relative frequency of driver mutations across early clonal and late clonal/subclonal stages, for the most common driver genes in HNC ($n$=173).

**Supplementary Figure 15. Human papillomavirus integration in HNC tumors. a,** Frequency of HPV16 integrations in genic and non-genic regions. **b,** Integration sites detected in chromosomal, cytoband,

and genic regions. Rows represent samples positive for viral integration. The number of integrations per site and sample is depicted. Only four samples presented integrations in genic regions. **c-d,** Circos plots representing viral integration sites, structural variations (SV) and copy number (CN) alterations in tumor genomes presenting HPV16 integration. HPV integrations (in yellow) are depicted in the outermost ring, CN in the inner ring, and SV events in the center. Specific SV and CN events surrounding the sites of integration (dotted lines) are shown for three samples (**c**).

**Supplementary Figure 16. Associations between germline ADH1B and ADH7 variant genotype and alcohol related mutational signatures.** *ADH1B* rs1229984 and *ADH7* rs1573496 germline variant genotypes for signatures SBS16 (a), DBS4 (b) and ID11 (c) (*n*=265 biologically independent samples). Mutated samples correspond to those with at least one alternative allele. The Kruskal–Wallis test (two sided) was used to test for global differences. Box-and-whisker plots are in the style of Tukey. The line within the box is plotted at the median, while upper and lower ends indicate 25th and 75th percentiles. Whiskers show 1.5 × interquartile range (IQR), and values outside it are shown as individual data points. Frequencies of positive samples in each category are indicated.

#### Supplementary Figure 1

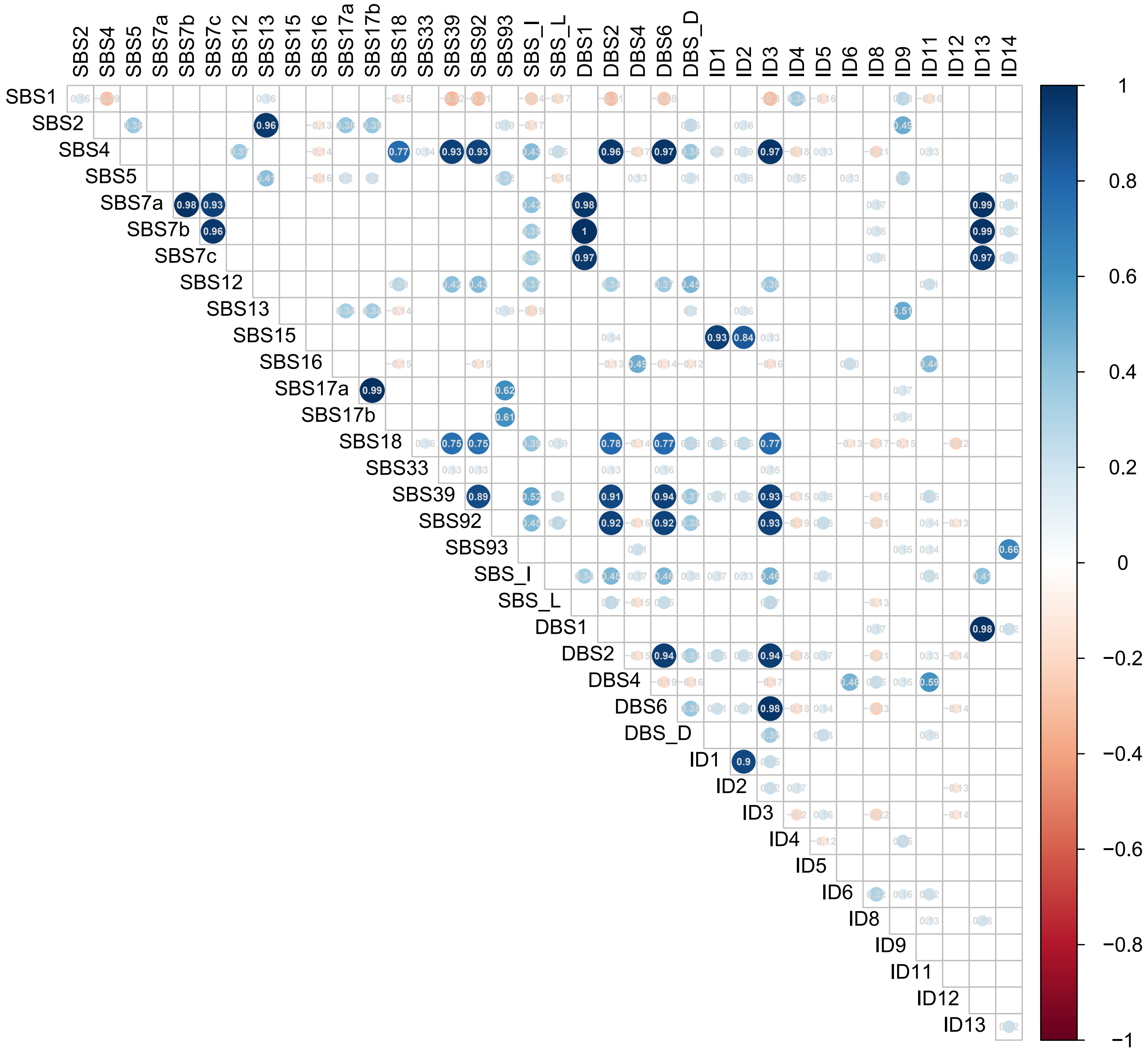

### Supplementary Figure 2

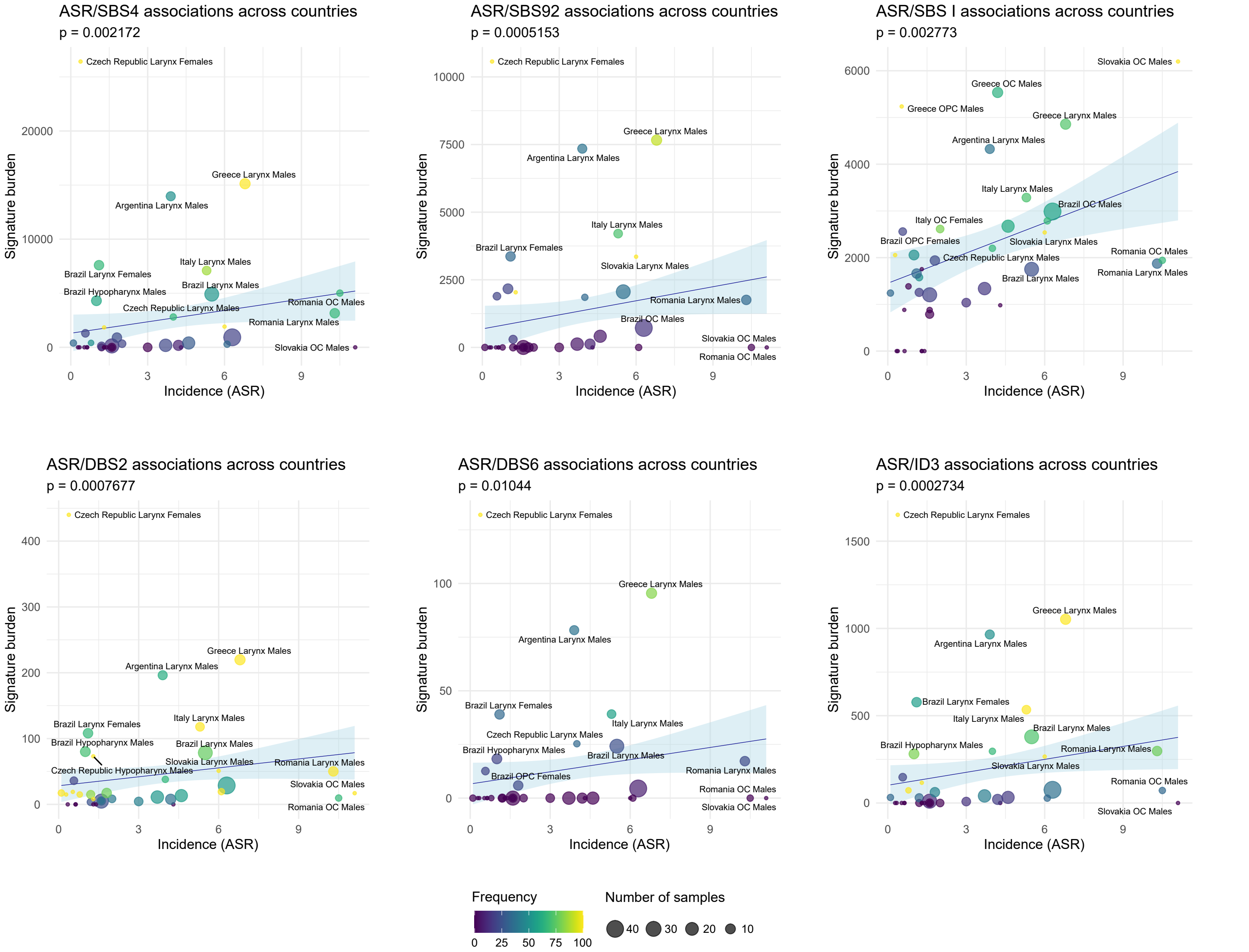

### Supplementary Figure 3

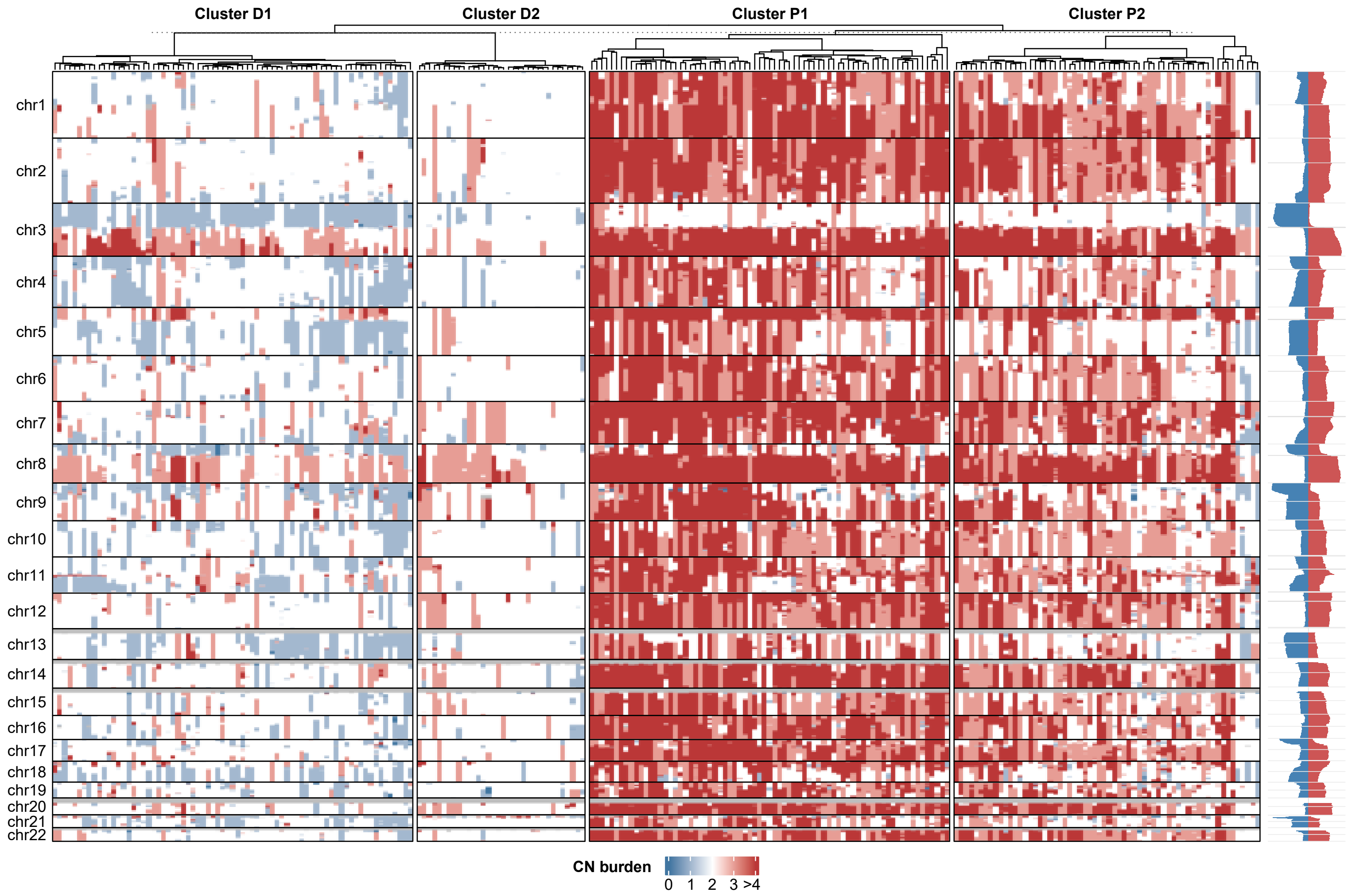

### Supplementary Figure 4

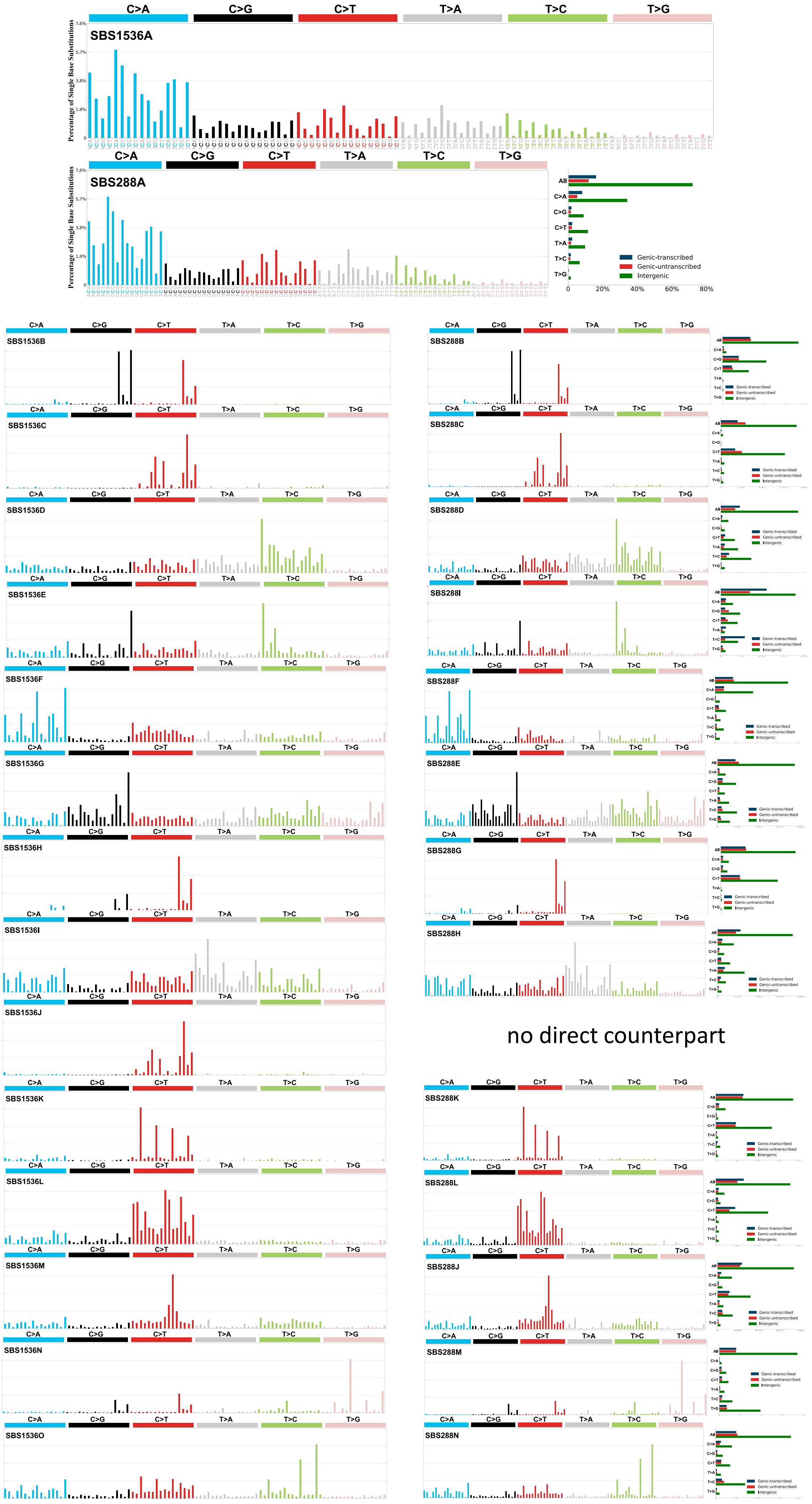

### Supplementary Figure 5

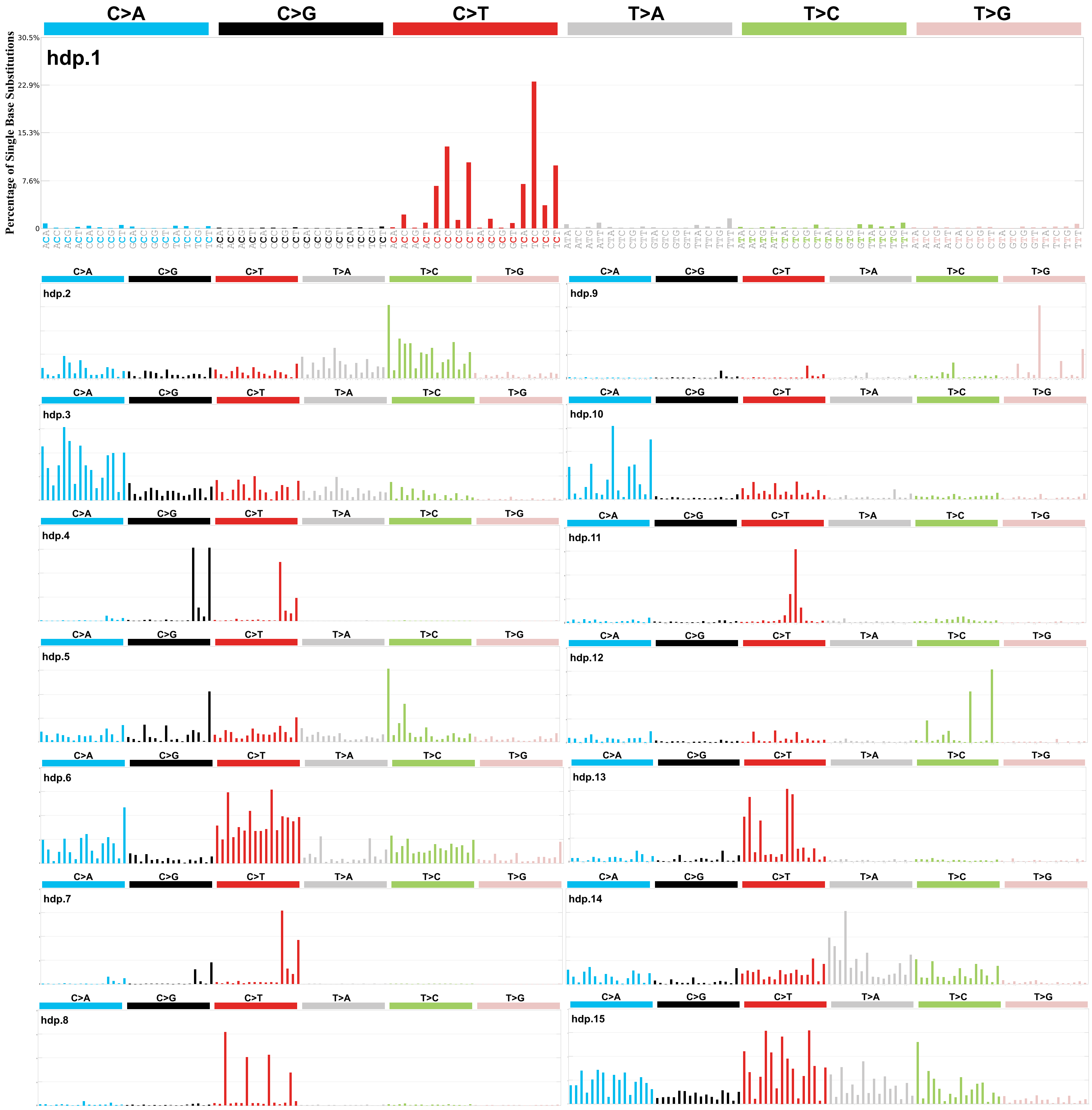

### Supplementary Figure 6

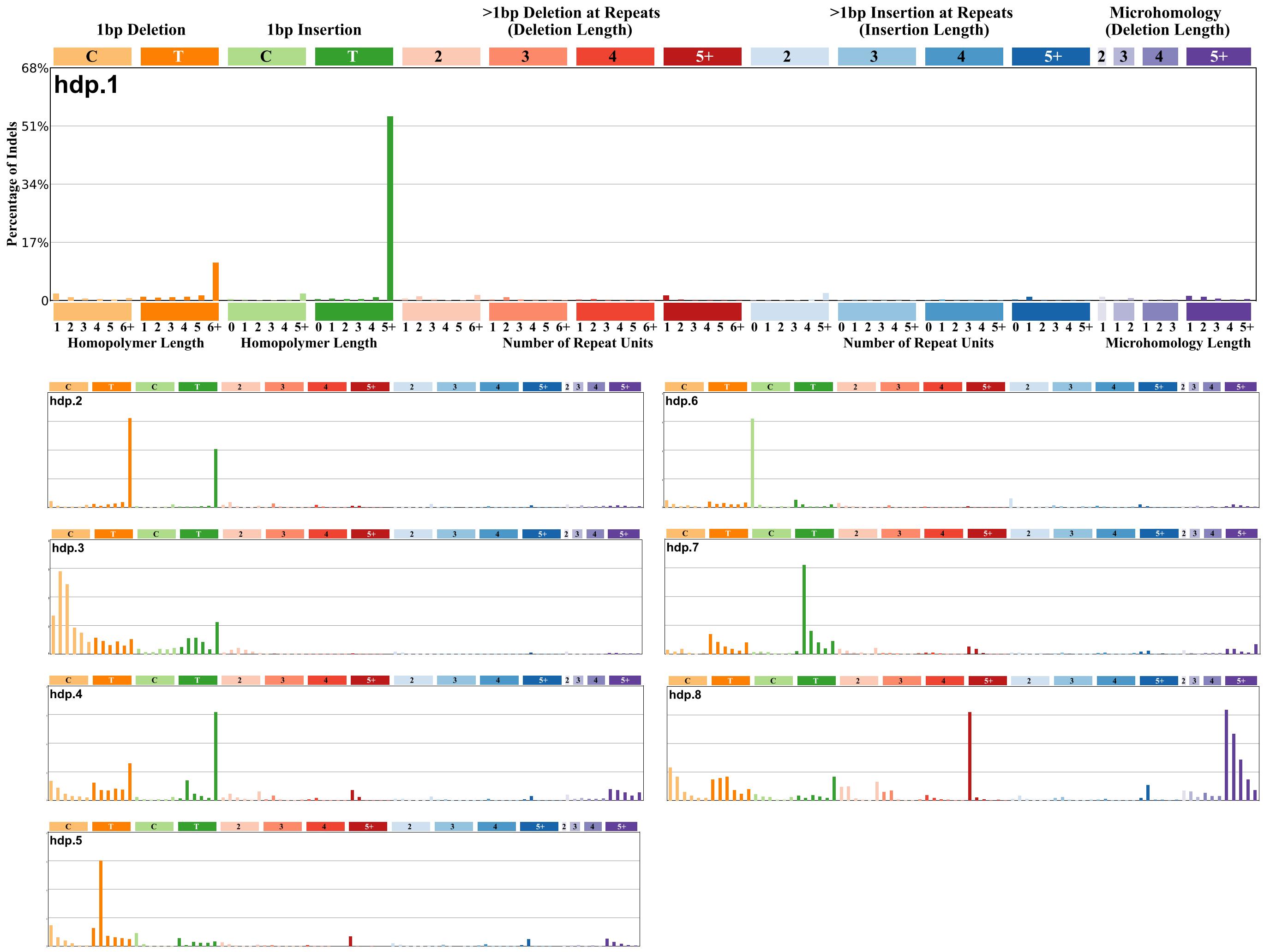

### Supplementary Figure 7

**a**

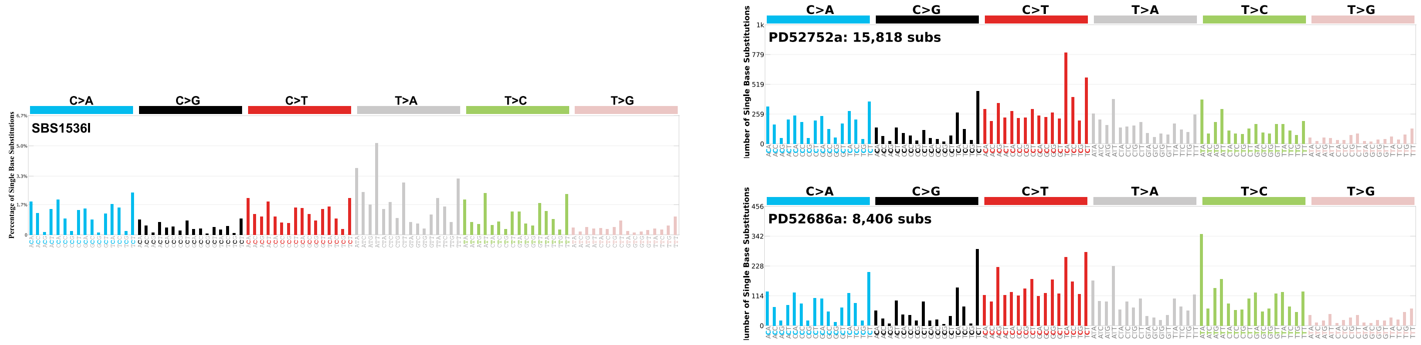


**b**

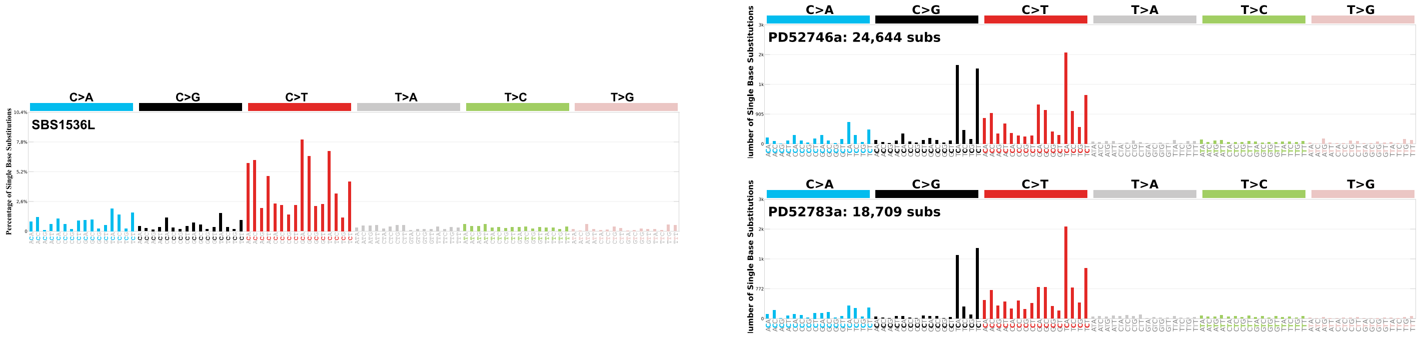


**c**

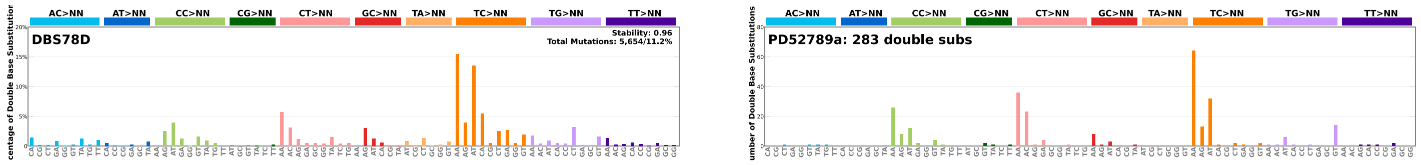

#### Supplementary Figure 8

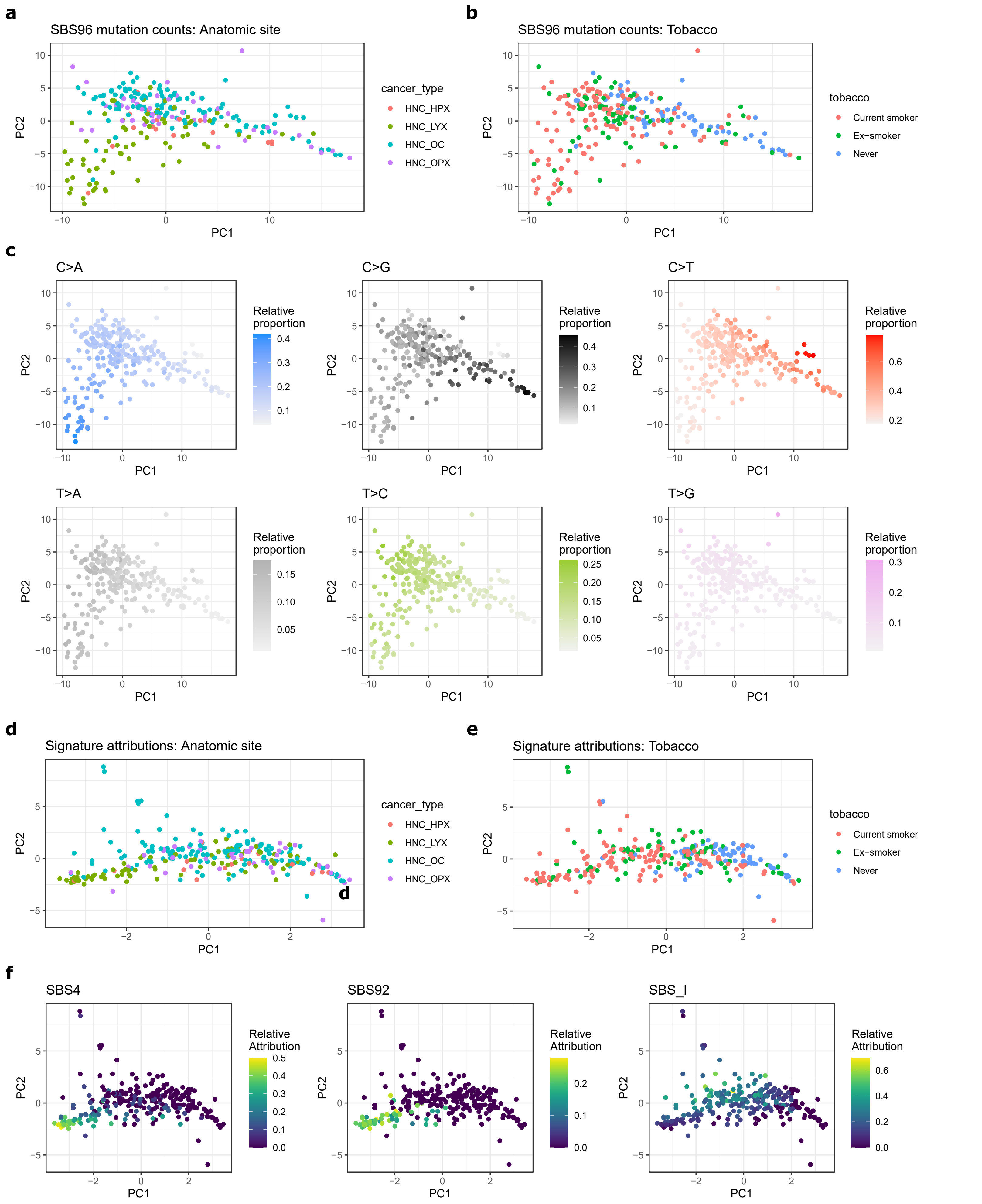

### Supplementary Figure 9

**a**

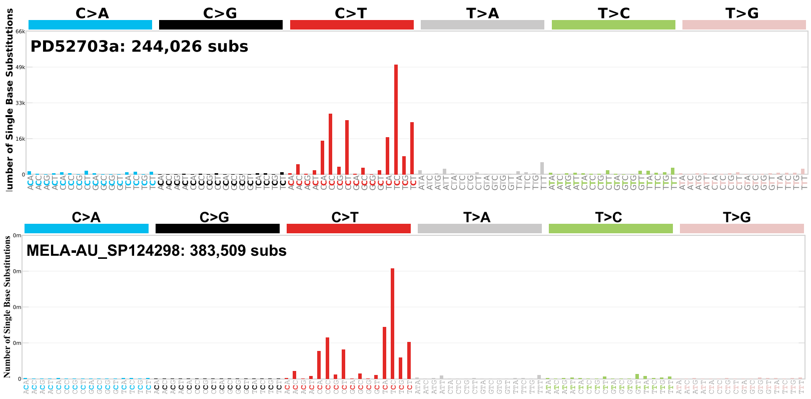


**b**

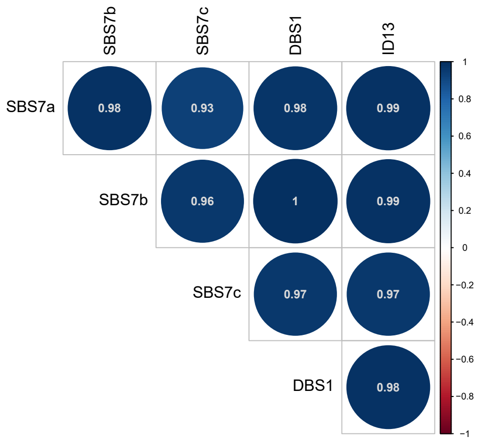

#### Supplementary Figure 10

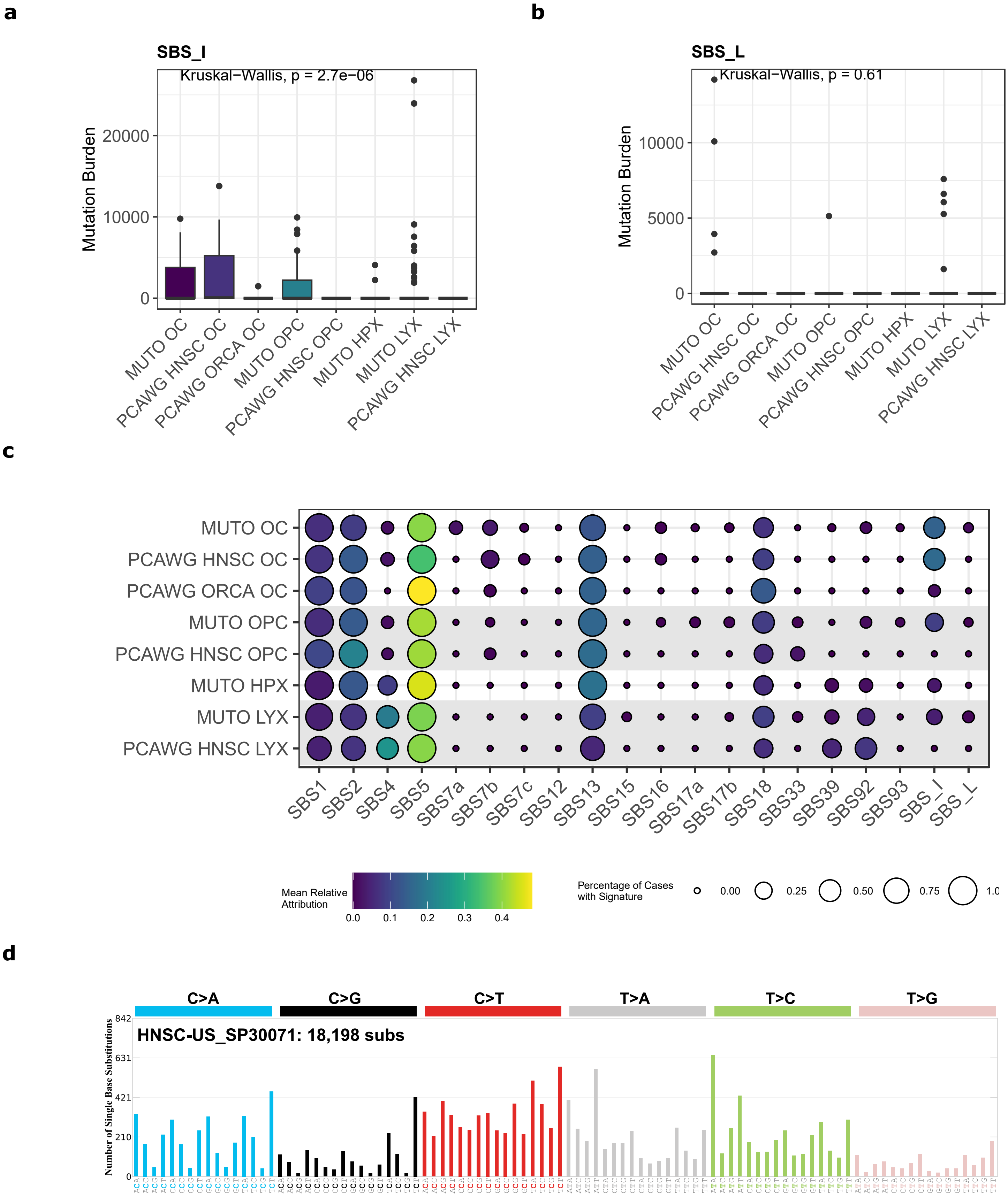

**Supplementary Figure 11**

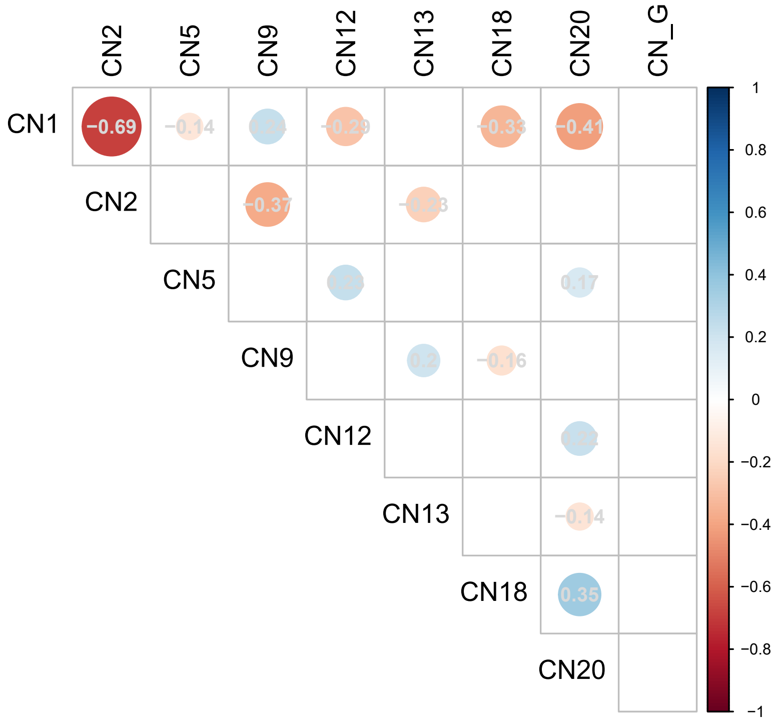

#### Supplementary Figure 12

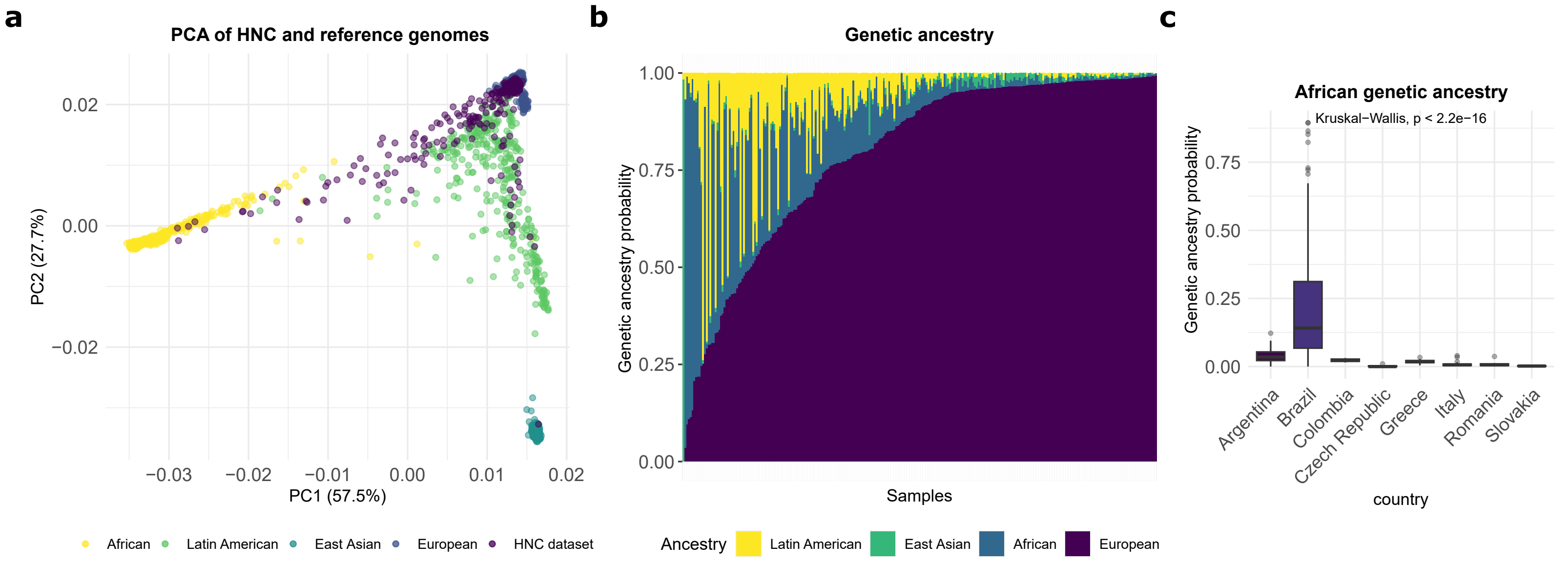

#### Supplementary Figure 13

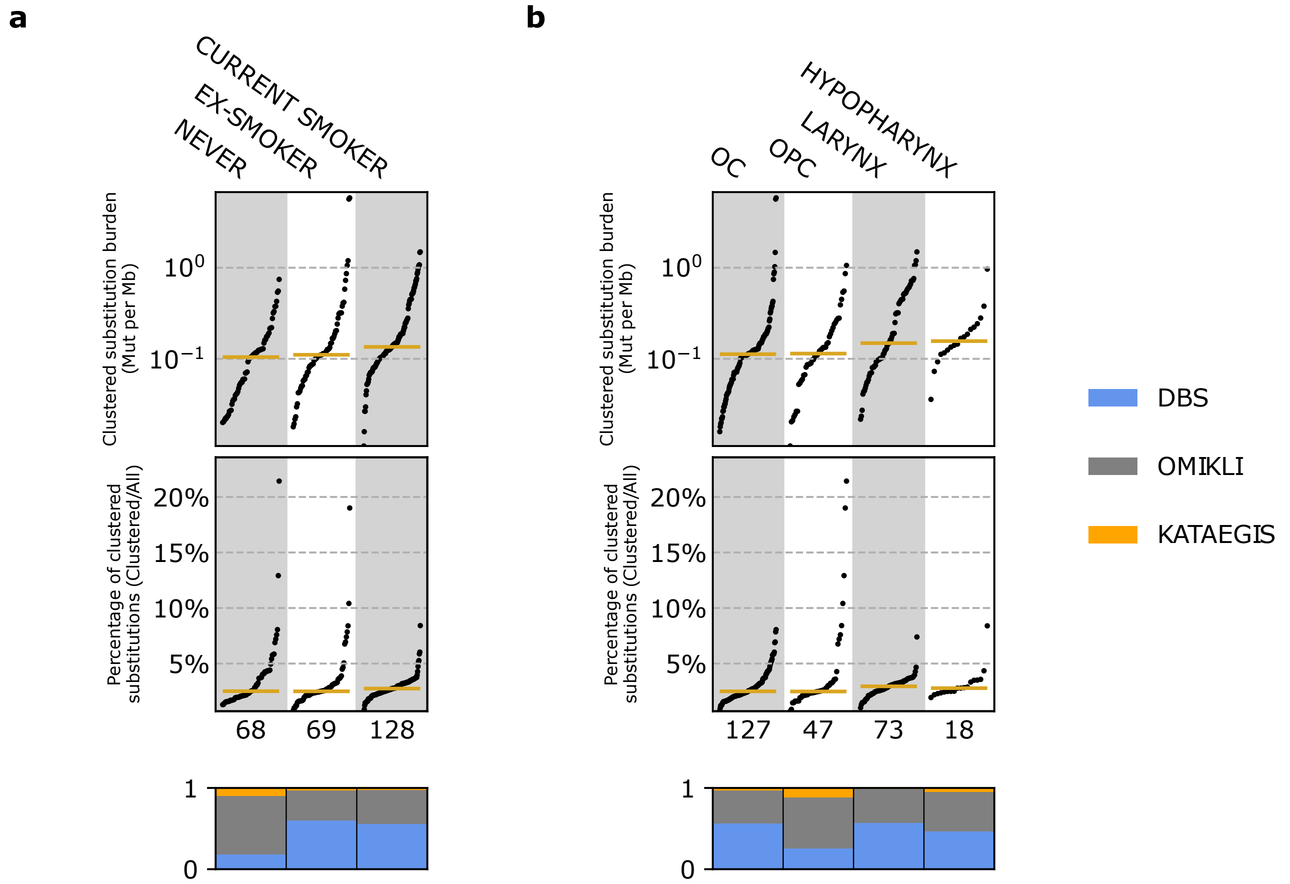

**Supplementary Figure 14**

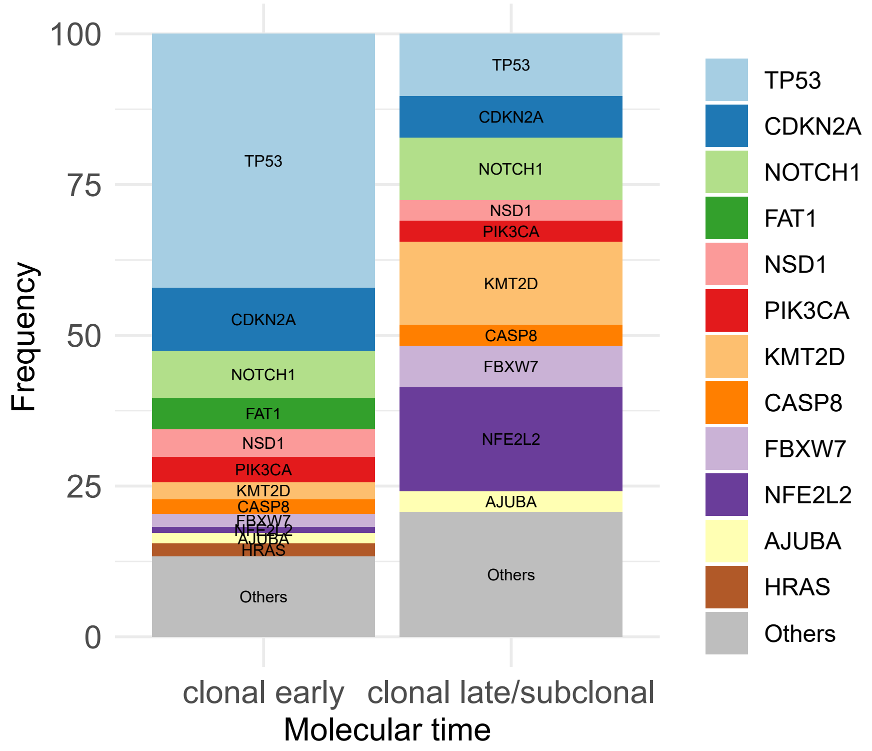

### Supplementary Figure 15

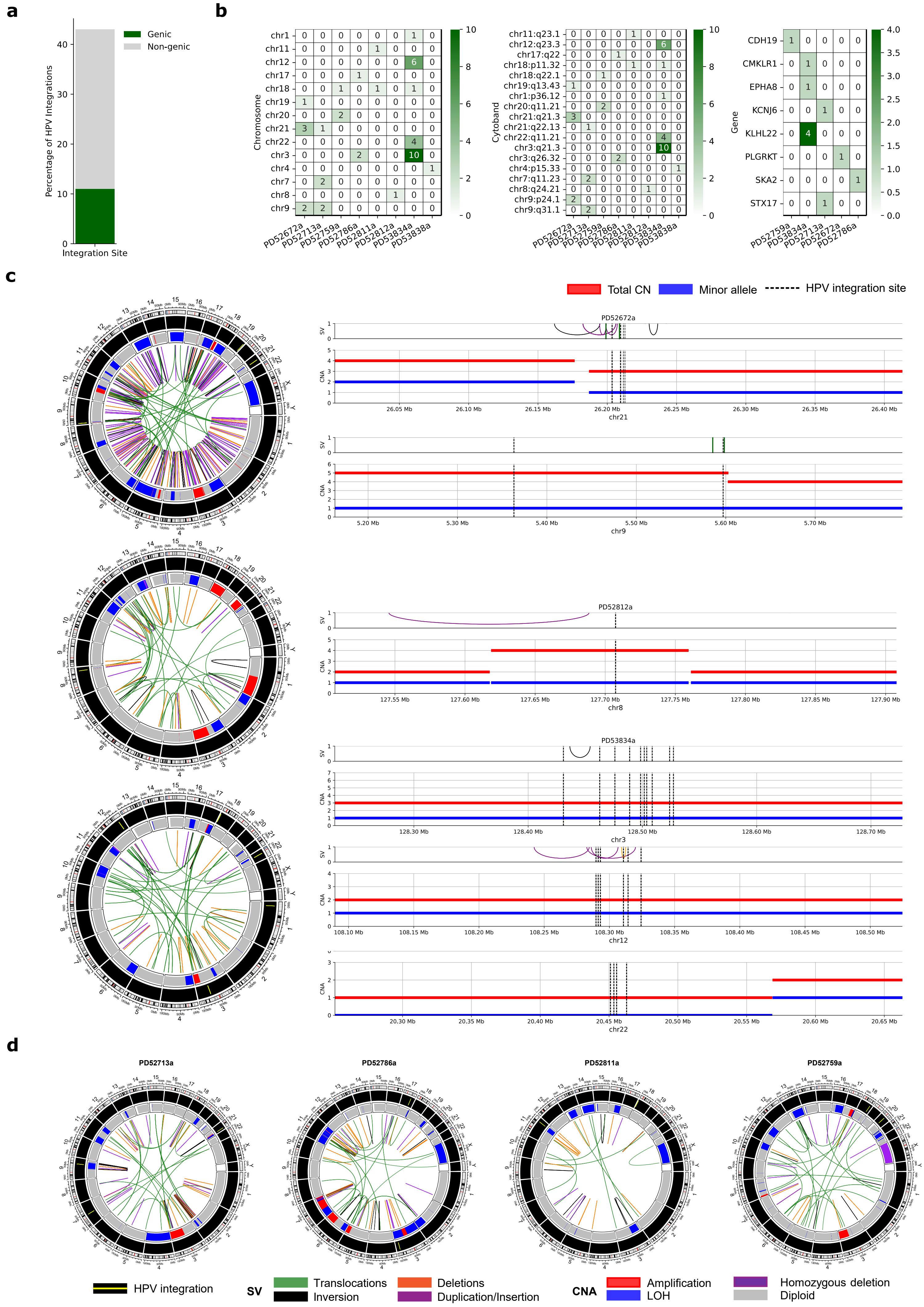

#### Supplementary Figure 16

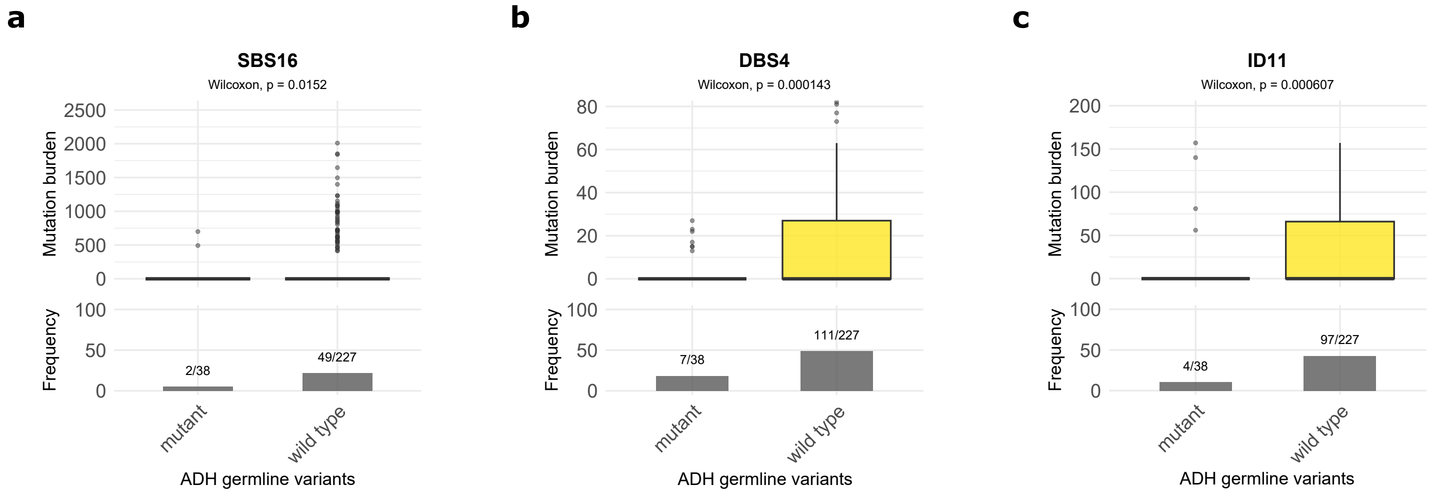
